# Genetic Associations of Lipid-Lowering Drugs and Parkinson’s Disease: A Mendelian Randomization Study

**DOI:** 10.1101/2024.01.14.24301288

**Authors:** Lin Shen, Yifan Yang, Yi Li, Hok Leong Chin, Helen Huang, Bernard Man Yung Cheung, Gary Tse, Oscar Hou In Chou, Jiandong Zhou

## Abstract

**Background:** There is a need to establish the role of lipid-lowering agents as a therapeutic option for Parkinson’s Disease (PD), but its associations remain elusive. This study investigated genetic variants proxying lipid-lowering agents through HMGCR, NPC1L1, and PCSK9 inhibitors to determine casual associations with PD risk.

**Methods:** We utilized a two-sample Mendelian randomization (MR) framework, where low-density lipoprotein (LDL) was the outcome of interest. Genetic associations with LDL were extracted from the Global Lipids Genetics Consortium. Summary statistics for PD were extracted from two GWAS datasets, consistent of 1,843 PD cases and 216,630 control in the first dataset and 1,570 PD cases and 1,259 controls in the second dataset. Instrumental variables (IV) were optimized with positive control analyses on cardiovascular and metabolic outcomes. IV-exposure associations from LDL GWAS data were integrated with IV-outcome associations from the PD GWAS data. The inverse variance weighted method was applied. Bayesian colocalization analysis identified target gene regions for LDL and PD.

**Results:** Genetic variations in *HMGCR* were significantly associated with a reduced risk of PD (odds ratio [OR] = 0.54, 95% CI 0.34-0.86). However, variation in *HMGCR* was associated with an increased risk of the tremor-dominant (TD) subtype compared to the postural instability/gait difficulty (PIGD) subtype (OR = 8.43, 95% CI 2.12-33.52). There were trends with increased risk for the TD subtype in *NPC1L1* and a decreased risk in *PCSK9* but these findings did not meet the Bonferroni threshold. We identified two single nucleotide polymorphisms (SNPs) in *HMGCR* within the same genomic region of close proximity, with rs12916 as the leading SNP associated with LDL and rs10942735 as the leading SNP associated with PD.

**Conclusion:** A casual association between *HMGCR* inhibition and reduced overall PD risk was identified, but there were increased the risks of tremor-dominant subtypes.

## Introduction

Parkinson disease (PD) is the second most common neurodegenerative disease with progressive nature^1^. In 2017, there were 1.02 million incident cases of PD as reported by the Global Burden of Disease^2^. The determinants of PD are multifactorial, with age-related susceptibilities being the biggest risk factor for PD with a median onset of 60 years of age^3^. Various studies have identified that genetic susceptibility could modify the effects of environmental stressors. In a recent meta-analysis of genome-wide association studies, 70 variants involved with the folding of alpha-synuclein may contribute to PD risk suggesting a genetic etiology^4–6^. Unfortunately, there are a lack of therapies that could delay the progression of disease.

With recent advancements in surrounding lipid-lowering therapies, targets of cholesterol metabolism may be of interest in PD. Lipids are becoming increasingly recognized to play a significant role in PD. Alpha synuclein is highly expressed in the brain, the second most lipid-rich organ, and exhibits binding properties with lipid membranes, thereby altering their structure and resulting in the formation of toxic alpha-synuclein oligomers^7,8^. There is much interest on how lipid aberrations could contribute to PD, as several risk genes exert known lipid-related functions^9^. An increase in serum lipids such as low-density lipoprotein (LDL) and its metabolites are correlated with oxidative stress in the brain through the generation of Lewy bodies, though there are conflicting studies suggesting that cholesterol and triglyceride levels are neuroprotective^10–13^. The role of lipid-lowering agents, such as 3-hydroxy-3-methylglutaryl-CoA reductase (HMGCR) inhibitors, Niemann-Pick C1-like 1 (NPC1L1) inhibitor, and proprotein convertase subtilisin/kexin type 9 (PCSK9) inhibitors, may be of therapeutic interest to ameliorate PD risk by targeting brain cholesterol metabolism and associated pathways but further studies are required to clarify these outcomes due to the bidirectional effect of lipids in PD^14^.

In this study, we leveraged genetic variants proxying LDL lowering agents through HMGCR, NPC1L1, and PCSK9 inhibitors to determine its association with PD. Genetic proxies have been successfully used to study the effect of lipid-lowering drugs which target alleles encoding various molecular stages in VLDL trafficking and cholesterol metabolism^15^. To establish casual associations of modifiable exposures such as genetic proxies for lipid-lowering therapies in PD, we utilized a two-sample Mendelian randomization (MR) framework to reduce reverse causation biases and less likely to be influenced by confounding factors^15^. This study aimed to investigate the relationship between lipid-lowering medication and PD.

## Methods

### Study Design

MR is a statistical technique that utilizes genetic variants as instrumental variables (IVs) to establish causal inferences between exposures and outcomes. This study employed the two sample MR framework to investigate the effects of genetic variation in lipid-lowering drug targets on the risk of Parkinson’s disease (PD)^16^. Exposure and outcome data were extracted from public genome wide association study (GWAS) summary datasets conducted on two independent, non-overlapping populations. risk of Parkinson’s disease. The details of the framework are presented in **eFigure 1**.

### LDL GWAS Data

Due to the unavailability of GWAS data on the exposure (protein targets of lipid-lowering drugs) and considering reducing LDL levels is a confirmed physiological response to lipid-lowering drug treatment, our study opts to use the available LDL as a biomarker of interest. This choice enables us to infer the effects of genetic variations in lipid-lowering drugs on the. Genetic associations with LDL were extracted from the largest GWAS dataset conducted by the Global Lipids Genetics Consortium, involving 1.3 million individuals of European ancestry. The details of primary study is provided in reference^17^.

### Parkinson’s Disease GWAS Data

The summary statistics for PD were extracted from the two GWAS datasets, which were published by the International Parkinson’s Disease Genomics Consortium. The first GWAS dataset consisted of 1,843 cases and 216,630 controls^6^. The other GWAS dataset focused on the motor subtype of PD consisted of 1,570 cases and 1,259 controls, where the tremor-dominant (TD) was defined as 1 and considered the case group, and the postural instability/gait difficulty (PIGD) was defined as 0 and served as the control group^18^. Both GWAS dataset were conducted on individuals of European ancestry to minimize potential bias from population stratification.

### Instrument Selection

The IVs in this study were screened for the genes that encode lipid-lowering drug target proteins. Variations within and around these genes were used to assess the effect of drug use on exposure factors and outcomes. Three major classes of lipid-lowering drugs were identified, including 3-hydroxy-3-methylglutaryl-CoA reductase (HMGCR) inhibitors, Niemann-Pick C1-like 1 (NPC1L1) inhibitors, and proprotein convertase subtilisin/kexin type 9 (PCSK9) inhibitors. Detailed information about the protein targets and encoding genes for these drugs is provided in **Table 1**.

**Table 1.** Detailed Information of Lipid-lowering Drug Classes, Targets, and Encodi ng Genes.

| Drug class | Representative drug | Drug target<br>(*Encoding gene*) | Function of target protein | Gene region |
| --- | --- | --- | --- | --- |
| 3-hydroxy-3-methylglutaryl-CoA reductase (HMGCR) inhibitors | Lovastatin | HMGCR<br>(*HMGCR*) | NADPH binding | Chr5:74632154-74657929 |
| Niemann-Pick C1-like 1 (NPC1L1) inhibitors | Ezetimibe | NPC1L1<br>(*NPC1L1*) | Rab GTPase binding | Chr7:44552134-44580914 |
| Proprotein convertase subtilisin/kexin type 9 (PCSK9) inhibitors | Alirocumab | PCSK9 (*PCSK9*) | Very-low-density lipoprotein particle receptor binding | Chr1:55505221-55530525 |

The single-nucleotide variations (SNVs) were designated within each encoding gene (within ±100kb base pairs of the gene location). Following this, variants with p-values greater than 5*10-8 and a minor allele frequency (MAF) below 0.01 werwe excluded in the GWAS data for LDL. The remained variants for each lipid-lowing drug target were selected as IVs and clumped with an R^2^ of 0.1, a window size of 10000 kb, and the reference population set to EUR. The detailed screening process of IVs for lipid-lowering drug is shown in **Figure 1**.

**Figure 1.**
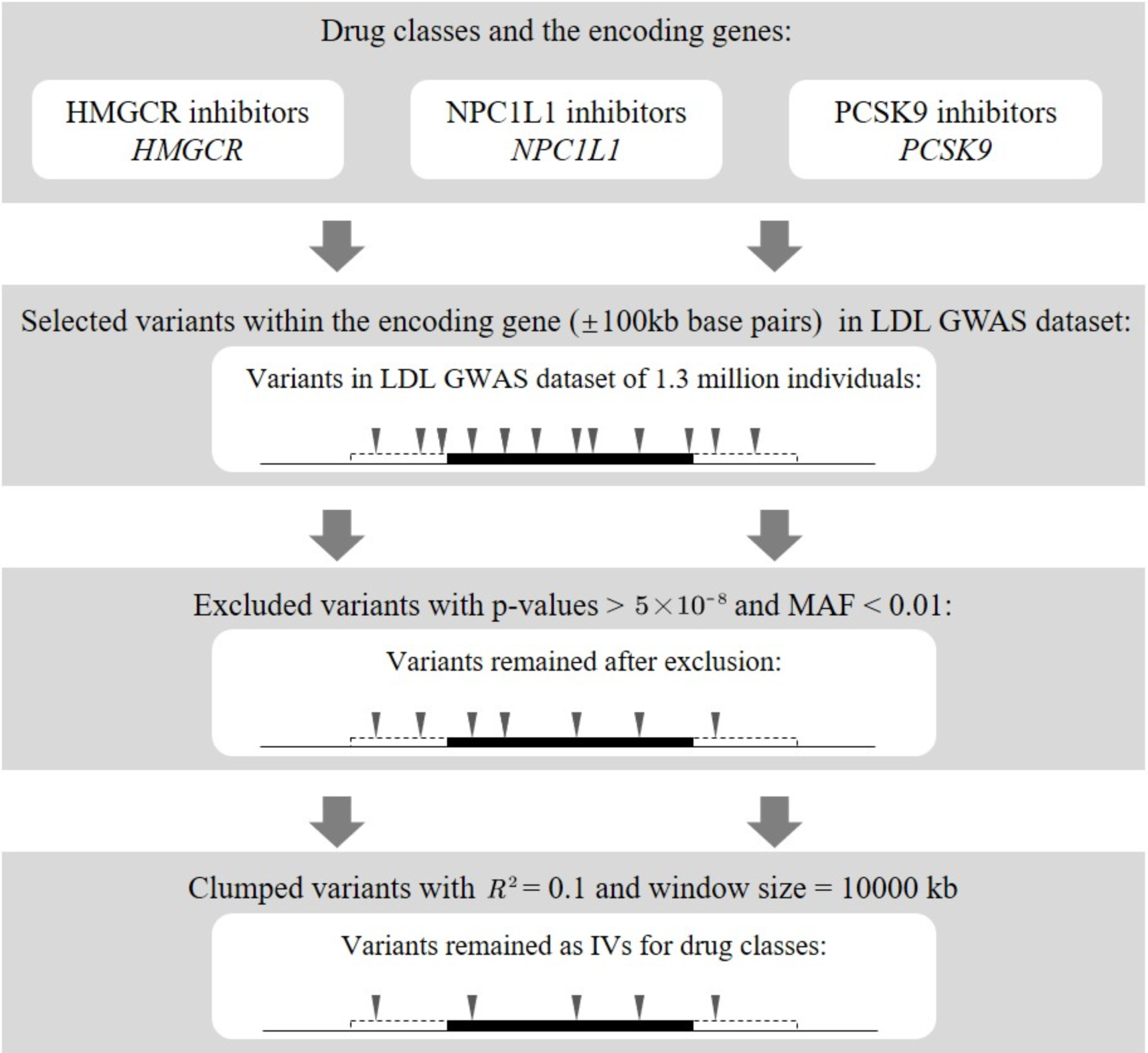
Instrument Selection for Lipid-lowering Drug Classes. The black line represents the DNA strand. The raised black box indicates the tar get gene, and the box with dashed line encompasses the region of ±100kb base pairs around the target gene. Each inverted triangle represents a genetic variant. Disappearing of inverted triangles indicates the exclusion of variants. HMGCR, 3-hydroxy-3-methylglutaryl-CoA reductase; MAF, minor allele frequency; NPC1L1, Niemann-Pick C1-like 1; PCSK9, proprotein convertase subtilisin/kexin type.

### Positive Control Analyses

In order to substantiate IVs selection, positive control analyses were performed using hyperlipidemia and coronary artery disease (CAD) as outcomes. Furthermore, three obesity-related traits were used as additional outcomes for validation, including body mass index (BMI), waist circumference (WC), and hip circumference (HC).

### Statistical Analysis

The present investigation integrated the IV-exposure association derived from the LDL GWAS data with the IV-outcome association obtained from the Parkinson’s disease GWAS data to create harmonized data for the MR study. To safeguard the validity of the IV, it is crucial to adhere to three fundamental assumptions. The initial assumption, known as relevance, necessitates that the genetic variants are significantly associated with the exposure. Evaluation of the strength of the IV was conducted through the computation of F statistics, derived as the square of the beta divided by the square of the standard error, with F statistics exceeding 10 indicatives of robust IV strength. The second assumption, independence, posits that the genetic variants remain unaffected by potential confounding factors, a condition typically fulfilled owing to the random assortment of genetic variants during conception. The third assumption, exclusion restriction, asserts that the variants exclusively influence the outcome through the designated exposure, thereby precluding horizontal pleiotropy. Evaluation of horizontal pleiotropy was conducted utilizing the MR Egger method, and non-significant results suggest the absence of horizontal pleiotropy. In instances where all three assumptions are met, the Inverse Variance Weighted (IVW) method is applied to yield unbiased causal estimates pertaining to the associations between exposures and outcomes.

The F-statistic was calculated by dividing the square of beta by the square of the standard error, where an F-statistic value greater than 10 indicating sufficient instrument strength. The Cochran Q test was employed to assess heterogeneity within IVs, and non-significant results signify the absence of heterogeneity. To mimic the LDL-lowering effects of lipid-lowering drugs, the MR estimates were standardized to a 1 mmol/L decrease in LDL. Following Bonferroni corrected principle, the significance level of multiple testing for 3 drugs classes was set as *p* value < 0.017 (0.05/3).

For the lipid-lowering drugs identified to be associated with Parkinson’s disease, Bayesian colocalization analysis was conducted specifically within the target gene region for LDL and Parkinson’s disease. Another sensitivity analysis was the leave-one-out method, which entails systematically excluding each variant within the drug’s target gene region and subsequently calculating the meta-effects of the remaining variants. The absence of statistically significant changes in MR estimates following the removal of each variant implies the robustness of the obtained results. All analyses were performed in R (version = 4.3.1), mainly utilizing the TwoSampleMR (version = 0.5.7) and coloc (version = 5.2.3) packages. This study follows the Strengthening the Reporting of Observational Studies in Epidemiology (STROBE) MR guidelines.

### Data Availability

All GWAS data analyzed in this study are publicly available. The LDL GWAS data is downloaded from the website of the Global Lipids Genetics Consortium (https://csg.sph.umich.edu/willer/public/glgc-lipids2021). The PD GWAS data is released in the resources page of International Parkinson’s Disease Genomics Consortium (https://www.pdgenetics.org/resources). The GWAS data for positive control analyses are available through the IEU Open-GWAS Project database (https://gwas.mrcieu.ac.uk/). The corresponding GWAS-ids are ebi-a-GCST90104006 for hyperlipidemia; ebi-a-GCST003116 for coronary artery disease; ieu-b-40 for BMI; ukb-b-9405 for WC and ukb-b-15590 for HC. All code for MR analyses is available in a GitHub repository.

## Results

### Instrument Selection

For the primary analysis, 22 variants were selected to proxy LDL lowering through inhibition of HMGCR (mean F-statistic of 194), 12 for NPC1L1 (mean F-statistic of 109), and 37 for PCSK9 (mean F-statistic of 286). Detailed data of the IVs to proxy lipid-lowering drugs are shown in **eTable 1**.

### Positive Control Analyses

As shown in **Figure 2**, genetic variations in *HMGCR*, *NPC1L1*, and *PCSK9* were significantly associated with a reduced risk of hyperlipidemia and CAD (**Figure 2, panels A and B**). For obesity-related traits (**Figure 2, panels C-E**), genetic variation in *HMGCR* was significantly correlated with increased BMI, WC, and HC. However, genetic variations in *NPC1L1* and *PCSK9* demonstrated no apparent correlation with BMI, WC, and HC. Positive control analysis data for each lipid-lowering drug class are available in **eTable 2-6**.

**Figure 2.**
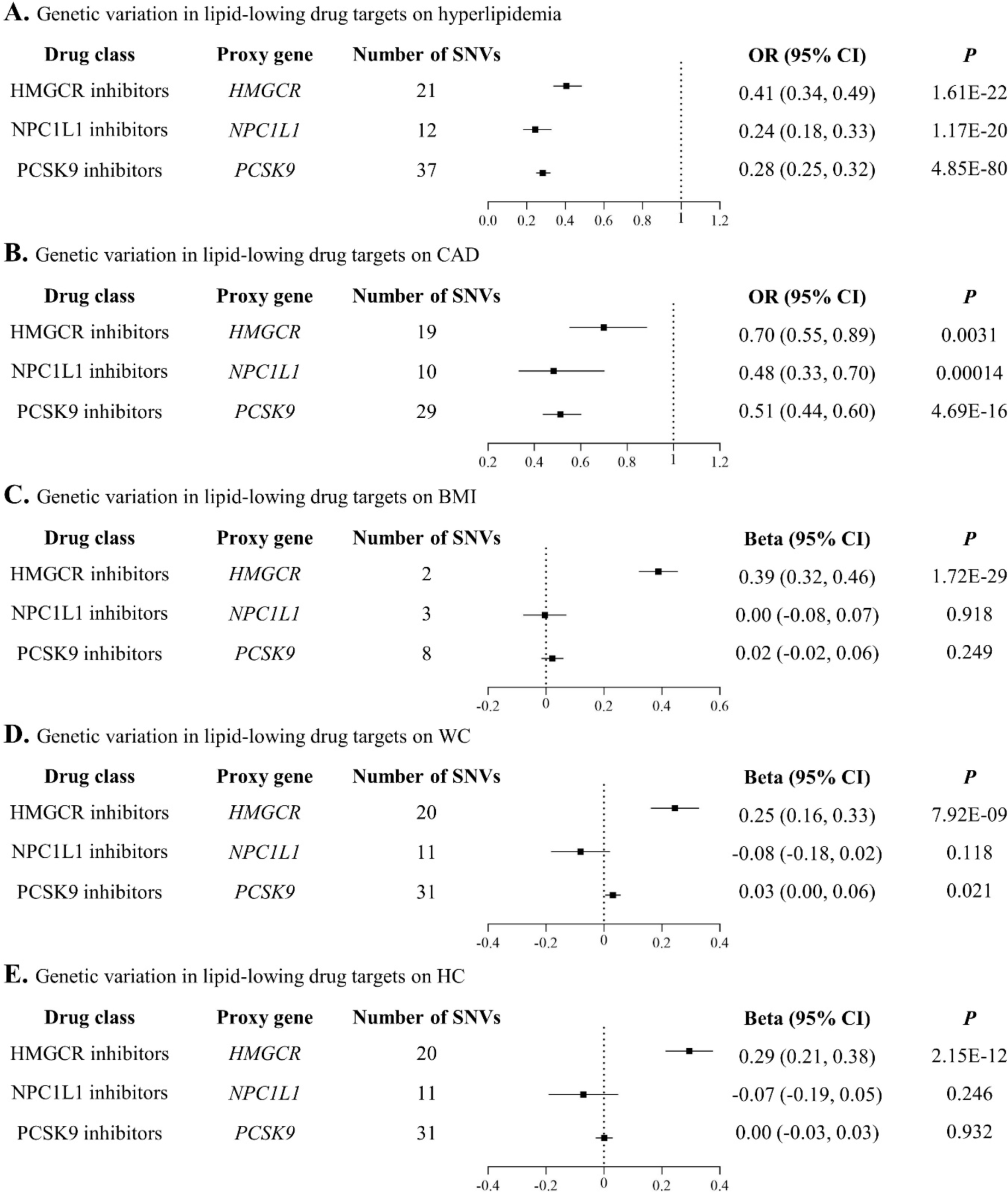
Estimated Effects of Genetic Variation in Lipid-lowing Drug Targets on Lipid Metabolism-Related Traits (Panel A-B) and Obesity-Related Traits (Panel C-E) Proxy gene is the gene that encodes the drug target protein. The estimated bet a coefficients on BMI, WC, and HC are expected to be in agreement with the e vidence from clinical trials, that is, taking in HMGB1 inhibitor (ie, lovastatin) cont ributes to weight gain, and taking NPC1L1 inhibitor (ie, ezetimibe) and PCSK9 in hibitor (ie, alirocumab) has no effect on weight. BMI, body mass index; CAD, coronary artery disease; HC, hip circumference; HMGCR, 3-hydroxy-3-methylglutaryl-CoA reductase; IVs, instrumental variables; NPC1L1, Niemann-Pick C1-like 1; OR, odds ratio; PCSK9, proprotein convertase subtilisin/kexin type; SNVs, single-nucleot ide variations; WC, waist circumference.

### Effects of Genetic Variation in lipid-lowering drug Targets on PD risk

We conducted MR analysis using two PD GWAS datasets, and the results are illustrated in **Figure 3**. For the unstratified PD dataset, the MR analysis results revealed a significant association between genetic variation in *HMGCR* and a reduced risk of PD at the Bonferroni-corrected threshold (odds ratio [OR] = 0.54, 95% CI 0.34-0.86, *p* = 0.009, **Figure 3A**). However, there was no apparent correlation between genetic variations in *NPC1L1* and *PCSK9* and the risk of PD.

**Figure 3.**
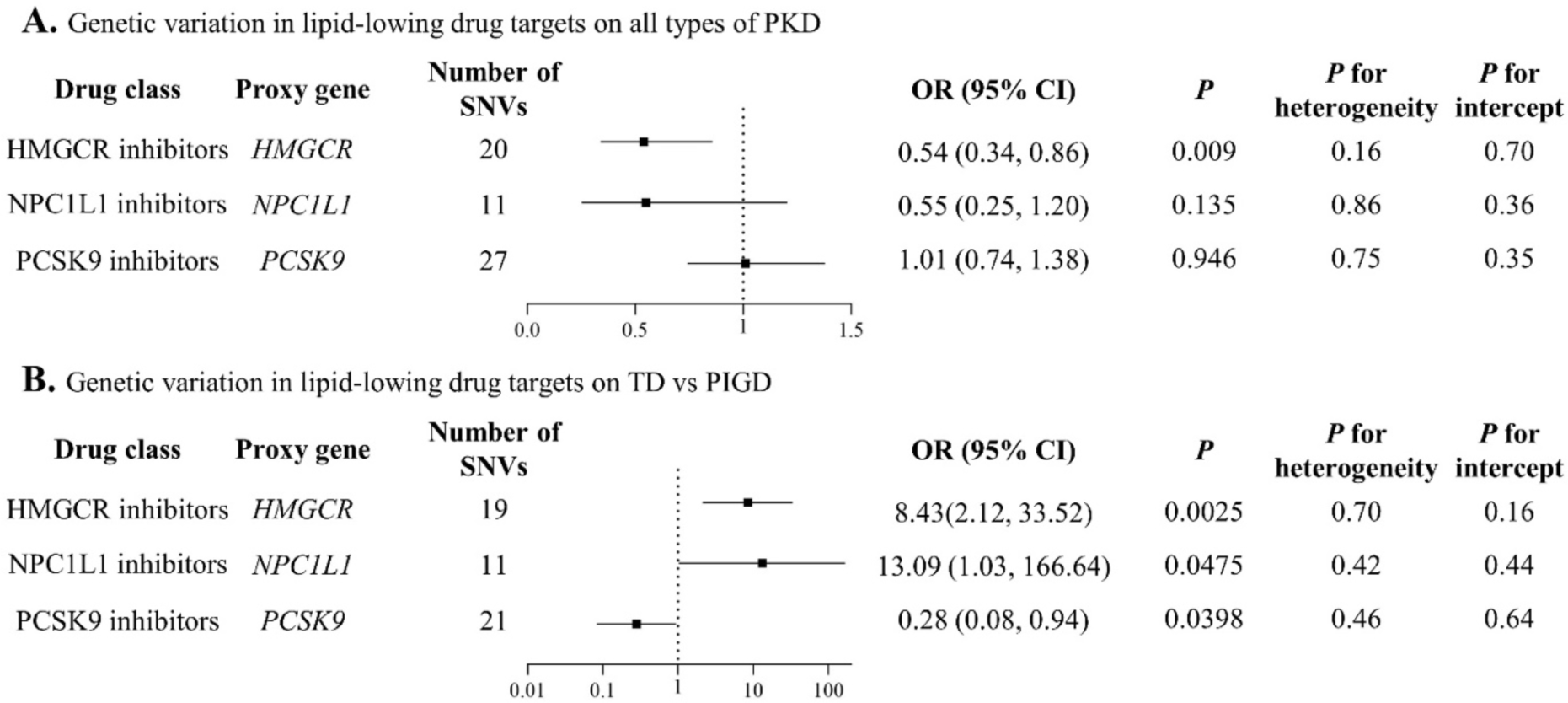
Estimated Effects of Genetic Variation in Lipid-lowing Drug Targets on Parkinson’s disease. Proxy gene is the gene that encodes the drug target proteins. *P* < 0.01 indicate s statistical significance, and 0.01 < *p* < 0.05 indicates suggestive significance. *P* for heterogeneity <0.05 indicates possible heterogeneity, whereas *p* for intercept <0.05 indicates substantial bias from pleiotropy. HMGCR, 3-hydroxy-3-methylglutaryl-CoA reductase; IVs, instrumental variables; MR, mendelian randomization; NPC1 L1, Niemann-Pick C1-like 1; OR, odds ratio; PCSK9, proprotein convertase subtilisin/kexin type; PIGD, postural instability/gait difficulty; PKD, Parkinson’s disease; SN Vs, single-nucleotide variations; TD, tremor-dominant.

Interestingly, further MR analysis on the PD dataset stratified by motor function showed that, compared to the PIGD subtype, genetic variation in *HMGCR* was significantly associated with an increased risk of the TD subtype at the Bonferroni-corrected threshold (OR = 8.43, 95% CI 2.12-33.52, *p* = 0.0025, **Figure 3B**). Additionally, genetic variation in *NPC1L1* showed a trend toward increased risk for the TD subtype (OR = 13.09, 95% CI 1.03-166.64, *p* = 0.0475), while genetic variation in *PCSK9* exhibited a trend toward decreased risk for the TD subtype (OR = 0.28, 95% CI 0.08-0.94, *p* = 0.0398). Unfortunately, the *p*-values for MR analysis of these two gene variations exceeded the Bonferroni-corrected threshold (*p* < 0.017).

As evidence suggested that cognitive function declines more rapidly, and prognosis is worse in PIGD patients compared to TD patients. Our analysis suggests that the use of HMGCR and NPC1L1 inhibitors may contribute to the transition of PD patients from PIGD subtype to TD subtype, while the use of PCSK9 inhibitors may pose a risk of worsening PD. No heterogeneity or pleiotropy was detected within the IVs in both PD GWAS datasets (**Figure 3**, *p*-values for heterogeneity > 0.05, *p*-values for pleiotropy > 0.05). Detailed MR analysis data are presented in **eTable 7-9**.

### Sensitivity Analysis

Colocalization analysis was conducted for LDL and PD within *HMGCR* (**eTable 10**), as well as co-localization analysis for LDL and PD motor subtype within both *HMGCR* and *PCSK9* (**eTable 11**). The results of the colocalization analysis indicated a low probability of shared random variation between LDL and PD within *HMGCR* (0.87%), and similarly low probabilities within both *HMGCR* and *PCSK9* for the motor subtype of PD (5.31% and 8.22%, respectively).

The findings suggest that there is no high probability of shared causal variants for LDL and PD (including the PD motor subtype) within the coding regions of *HMGCR* and *PCSK9*. However, in the regional plot illustrating the associations of LDL and PD within ±100kb of *HMGCR*, rs12916 emerged as the leading SNP associated with LDL, while rs10942735 was identified as the leading SNP associated with PD within the same genomic region. Notably, these two leading SNPs were in close proximity, that revealed a trend of colocalization between LDL and PD (**eFigure 2**).

Additionally, in the PD GWAS datasets, leave-one-out sensitivity analyses were conducted for the genetic variations in *HMGCR* and *PCSK9*. The analyses demonstrated the robustness of the results, indicating that after excluding each SNP, the overall error lines show minimal changes (all error lines are either to the right or left of 0), as shown in **eFigure 5-7**.

## Discussion

This study investigated the effects of genetic variations in lipid-lowering drugs on the PD risk using a combination of LDL genetic dataset in ∼1,300,000 participants and two PD genetic datasets, consistent of 1,843 PD cases and 216,630 control in the first dataset and 1,570 PD cases and 1,259 controls in the second dataset. To the best of our knowledge, this is one of the largest studies with strong validation suggesting a causal association between HMGCR inhibition and a reduced risk of PD. Besides, this study was the first MR to investigate the association between genetic variation in lipid-lowering drug and the subtypes of PD.

A handful of literature has explored the association between lipid-lowering drugs or serum lipid levels and PD, yet the results have been inconsistent ^19–26^. In a MR study using a Danish dataset, no association was detected between LDL and the risk of PD, whereas a separate MR study conducted by a Swedish team suggested a causal inverse association of LDL with PD risk^19,27^. Another recent MR study did not project an elevated risk of PD with statin exposure^20^. However, the results lacked sufficient precision to conclusively support preventive benefits. Nevertheless, this does not negate the potential advantages of on-target effects associated with statin use for PD prevention through alternative mechanisms^20^. However, all of these MR studies have their own statistical limitations. Other non-MR studies have also reported varied conclusions^21–25^. Interpreting the above studies alongside our study results, which showed that only HMGCR inhibition, not NPC1L inhibition and PCSK9 inhibition, was correlated with a reduced risk of PD. The result provided evidence that HMGCR inhibitors may modify the pathogenesis of PD through a mechanism different from lowering LDL. Indeed, multiple studies have proposed the protective effects of HMGCR inhibitors on α-synuclein aggregation through mechanisms such as antioxidation and promotion of neural repair^28–30^. Other suggested mechanisms involve HMGCR inhibitors preventing neuronal injury by mitigating microglia and autophagy activation^31–33^. As HMGCR inhibitors can cause a dose-dependent reduction in coenzyme Q10, the protective effects of HMGCR inhibitors may be compromised by mitochondrial dysfunction if the dose surpasses a specific threshold^34^. Nonetheless, this potential explanation for the inconsistency in previous studies warrants further investigation.

Another discovery from this study implied that incorporating HMGCR inhibitors may facilitate the shift of PD patients from the less favorable PIGD subtype to the more favorable TD subtype. The mechanisms behind the observations in the current study were not fully understood and necessitate additional research. However, previous studies have pointed to the involvement of various neural pathways in contributing to the PIGD phenotype of Parkinson’s disease^35–37^. For example, some literature hinted at a connection between the severity of PIGD and amyloid β pathology^35,38^. The neuroprotective impact of HMGCR inhibitors on amyloid β-induced neurotoxicity may offer a partial explanation for the effect observed on the PIGD subtype^39^. Other studies suggested that elevated GABA levels or reduced acetylcholine may play a role in the development of the PIGD phenotype in Parkinson’s disease, and HMGCR inhibitors might be involved in modifying the pathogenesis^40,41^. Given functional network alterations within cerebro-cerebellar motor pathways were identified among PD motor subtypes, we contemplate whether HMGCR inhibitors contribute to the alterations, warranting further exploration in future studies^37^.

This study possesses several strengths. First, it involves large sample sizes from public GWAS datasets for analysis. The inclusion of such substantial and robust data enables study replication and diminishes random errors. In addition, all the GWAS datasets used were conducted on individuals of European ancestry, thereby minimizing potential biases arising from population stratification. Secondly, the two-sample MR study design may overcome the challenges of confounding between the exposure and outcome when estimating the causal exposure-outcome association. It also offers an opportunity to enhance statistical power and avoid reverse causality. Third, by performing a series of positive control analyses and sensitivity analyses that considered different patterns of pleiotropy, this study enhanced the robustness of the results, thereby reinforcing the evidence supporting our findings.

The study has several limitations. First, MR study needed to fulfill the instrumental variable assumptions which cannot be empirically verified. Even though rigorous statistical methods were employed, it is important to note that the complete exclusion of pleiotropy and bias effects cannot be guaranteed. Second, the lipid-lowering drug effect predicted by MR cannot be directly juxtaposed with the pharmacological effect observed in real life practice. The exposure instrumented by genetic variants persists throughout lifetime, whereas pharmacological intervention influences only a specific period of life. Consequently, the results of this study should be interpreted as evaluating the long-term modulation effects of drug target proteins. Third, lipid-lowering drugs may modify PD through interactions with other protein targets not captured in this MR study or via different mechanisms. Fourthly, the dataset utilized in this study only comprised individuals of European ancestry, limiting the generalizability of the results to other ethnic groups. However causal effects are expected to be consistent across settings unless the mediating mechanisms differ, although the extent of response may vary.

## Conclusion

The two way Mendelian Randomization result identified the casual association between genetic variations in *HMGCR* and PD, which suggested that lipid-lowering drugs may play a role in lowering risks of PD. However, the risk of TD subtypes are higher compared to PIGD subtypes, suggesting the bidirectional effects of the lipid-lowering drugs.

## Ethical approval statement

None.

## Availability of data and materials

Data are publicly available and can be accessed with the GWASID.

## Supporting information

STROBE_checklist

## Data Availability

Data are publicly available and can be accessed with the GWASID.

## Acknowledgements

None

## Guarantor Statement

All authors approved the final version of the manuscript.

## Author contributions

L S, YF, JD Z: conception of study, preparation of figures, study design, data contribution, statistical analysis, data interpretation, manuscript drafting, and critical revision of the manuscript.

Y L, HL C, H H: literature search, data interpretation, literature search, data collection, manuscript drafting.

BMY C, G T: critical revision of the manuscript.

OHI C, JD Z: conception of study and literature search, study design, data collection, and critical revision of manuscript, study supervision.

