## Supplementary Appendix for "Genetic Associations of Lipid-Lowering Drugs and Parkinson’s Disease: A Mendelian Randomization Study"

Content outline:

**eFigure 1.** Framework for the 2-sample MR study design

**eFigure 2.** Regional plot for the associations of LDL and Parkinson's disease within ±100kb of *HMGCR*

**eFigure 3.** Regional plot for the associations of LDL and PKD subtype (TD vs PIGD) within ±100kb of *HMGCR*

**eFigure 4.** Regional plot for the associations of LDL and PKD subtype (TD vs PIGD) within ±100kb of *PCSK9*

**eFigure 5.** Leave-one-out sensitivity analysis for genetically proxied HMGCR inhibition on PKD

**eFigure 6.** Leave-one-out sensitivity analysis for genetically proxied HMGCR inhibition on PKD subtype (TD vs PIGD)

**eFigure 7.** Leave-one-out sensitivity analysis for genetically proxied PCSK9 inhibition on PKD subtype (TD vs PIGD)

**eTable 1.** Genetic variants used to instrument each lipid-lowering drug for the primary analysis

**eTable 2.** Characteristics of instrumental variables for each drug target in the LDL and hyperlipidemia datasets

**eTable 3.** Characteristics of instrumental variables for each drug target in the LDL and coronary artery disease datasets

**eTable 4.** Characteristics of instrumental variables for each drug target in the LDL and body mass index datasets

**eTable 5.** Characteristics of instrumental variables for each drug target in the LDL and waist circumference datasets

**eTable 6.** Characteristics of instrumental variables for each drug target in the LDL and Hip circumference datasets

**eTable 7.** Characteristics of instrumental variables for HMGCR protein target in the LDL and Parkinson's disease datasets

**eTable 8.** Characteristics of instrumental variables for NPC1L1 protein target in the LDL and Parkinson's disease datasets

**eTable 9.** Characteristics of instrumental variables for PCSK9 protein target in the LDL and Parkinson's disease datasets

**eTable 10.** Colocalization results in the target gene region for LDL and PKD

**eTable 11.** Colocalization results in the target gene region for LDL and PKD subtype (TD vs PIGD)

**eFigure 1. Framework for the 2-sample MR study design.**

This study employs the 2-sample MR to investigate the effects of genetic variation in lipid-lowering drug targets on the risk of Parkinson's disease (PKD). Specifically, the genetic variants (IVs) influences the risk of PKD (outcome) either directly or indirectly through its effect on the protein targets of lipid-lowering drugs (exposure). These drugs operate through low density lipoprotein (LDL, biomarker) levels, subsequently affect the risk of PKD. The genetic effects of this framework, labelled as the path, involving: the effect of IVs on exposure ($\delta$); the effect of IVs on confounders ($\rho_G$); the effect of IVs on biomarker ($\delta\mu$); the effect of IVs on outcome ($\phi_G + \delta(\phi_P + \mu\theta)$). To safeguard the validity of the IV, it is crucial to adhere to three fundamental assumptions, which are relevance ($\delta \neq 0$); independence ($\rho_G = 0$) and exclusion restriction ($\phi_G = 0$). In instances where all three assumptions are met, the associations between exposures and outcomes can be calculated:

$$\omega = \frac{\delta(\phi_P + \mu\theta)}{\delta} = \phi_P + \mu\theta.$$

Due to the unavailability of GWAS data on the exposure, we alternatively calculates the associations between biomarker and outcomes:

$$\omega_b = \frac{\delta(\phi_P + \mu\theta)}{\delta\mu} = \frac{\phi_P + \mu\theta}{\mu} = \frac{1}{\mu} \times \omega.$$

It is noteworthy that, despite $\omega_b$ not being equal to $\omega$, it can still provide a valid hypothesis test for $\omega = 0$, as exposure is significantly correlated with the biomarker.

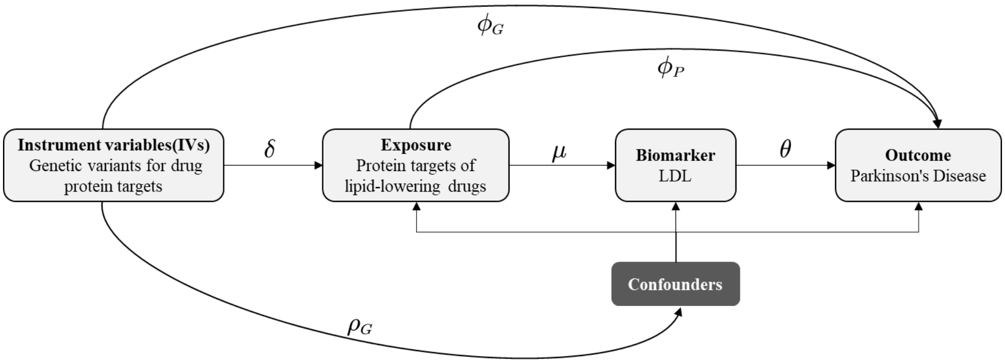

**eFigure 2. Regional plot for the associations of LDL and Parkinson's disease within ±100kb of *HMGCR*.**

*rs12916* is the leading SNP identified to be associated with LDL within ±100kb of *HMGCR* and colored as purple in the plot. *rs10942735* is the leading SNP identified to be associated with Parkinson's disease within the region and colored as purple in the plot. LDL, low density lipoprotein; SNP, single nucleotide polymorphism.

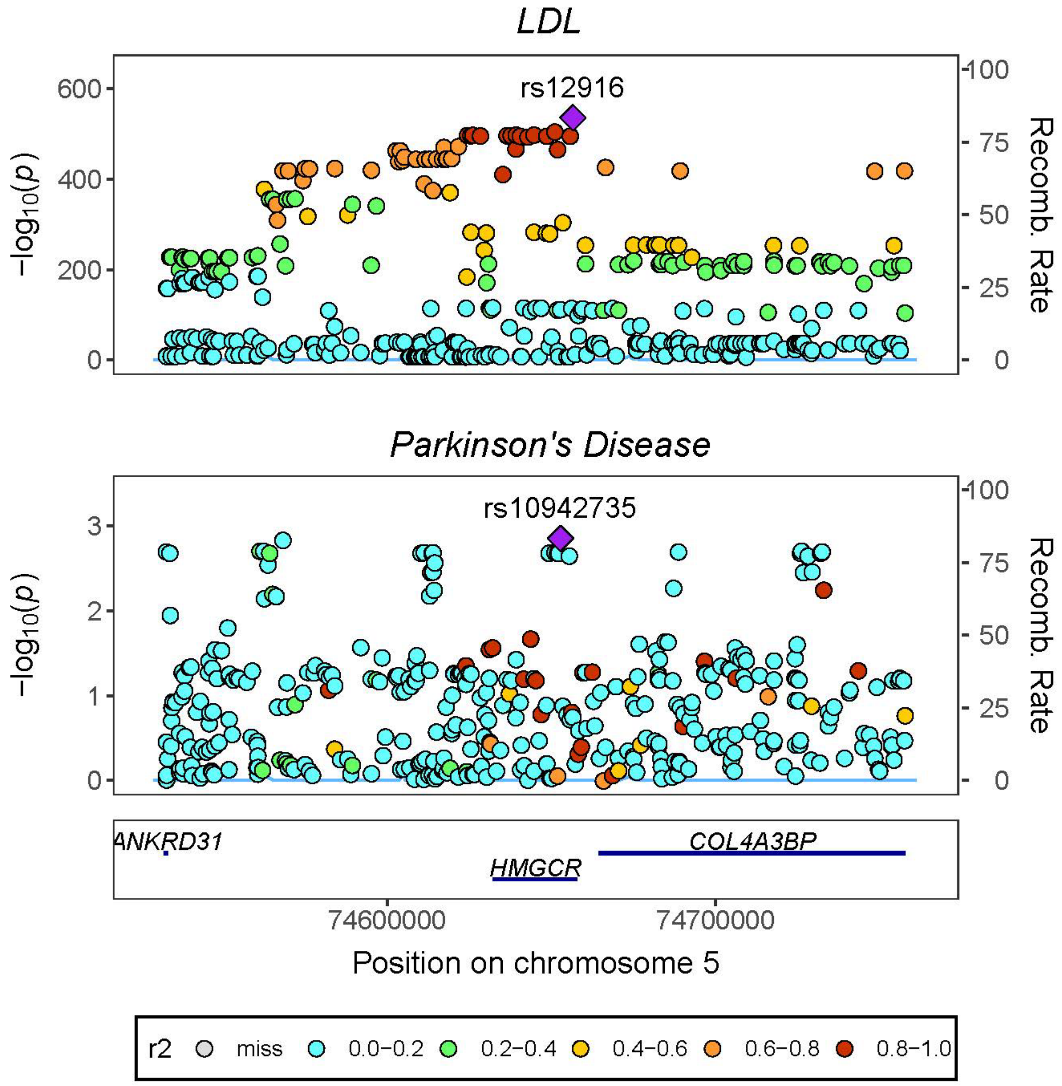

**eFigure 3. Regional plot for the associations of LDL and PKD subtype (TD vs PIGD) within ±100kb of *HMGCR*.**

*rs12916* is the leading SNP identified to be associated with LDL within ±100kb of *HMGCR* and colored as purple in the plot. *rs77096078* is the leading SNP identified to be associated with PKD subtype within the region and colored as purple in the plot. LDL, low density lipoprotein; PIGD, postural instability/gait difficulty; PKD, Parkinson's disease; TD, tremor-dominant.

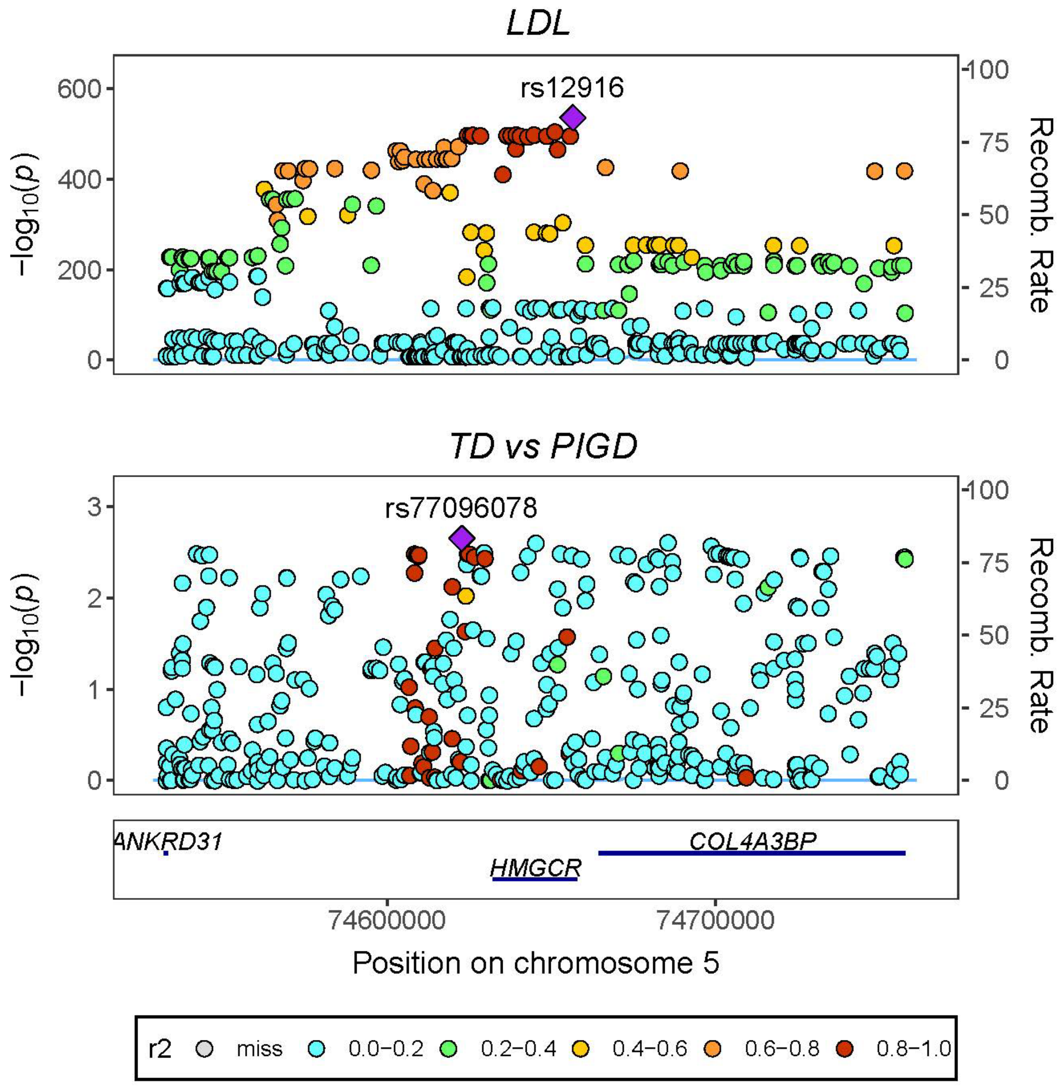

**eFigure 4. Regional plot for the associations of LDL and PKD subtype (TD vs PIGD) within ±100kb of *PCSK9*.**

*rs11591147* is the leading SNP identified to be associated with LDL within ±100kb of *PCSK9* and colored as purple in the plot. *rs11800231* is the leading SNP identified to be associated with PKD subtype within the region and colored as purple in the plot. LDL, low density lipoprotein; PIGD, postural instability/gait difficulty; PKD, Parkinson's disease; TD, tremor-dominant.

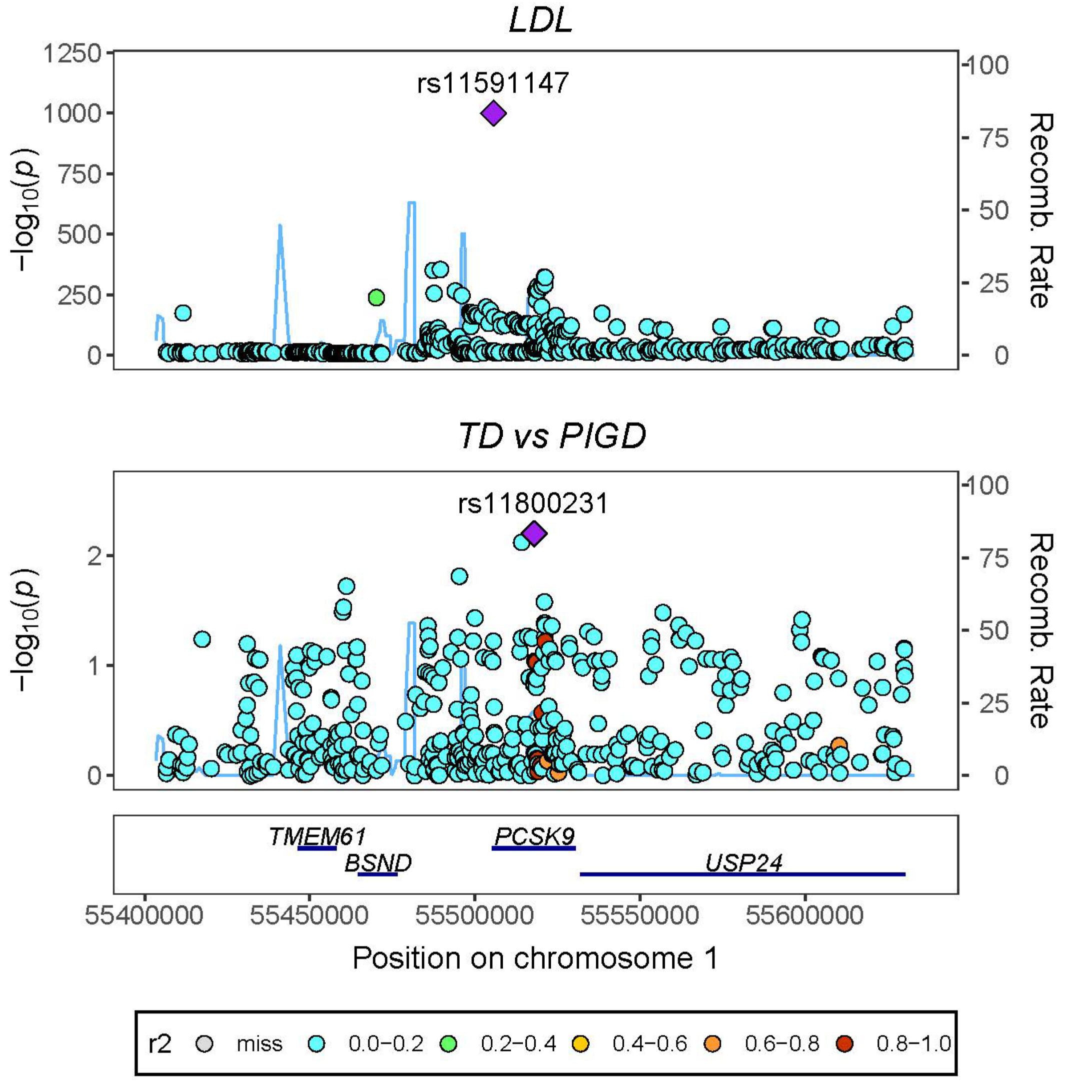

**eFigure 5. Leave-one-out sensitivity analysis for genetically proxied HMGCR in hibition on PKD.**

HMGCR, 3-hydroxy-3-methylglutaryl-CoA reductase; PKD, Parkinson's disease.

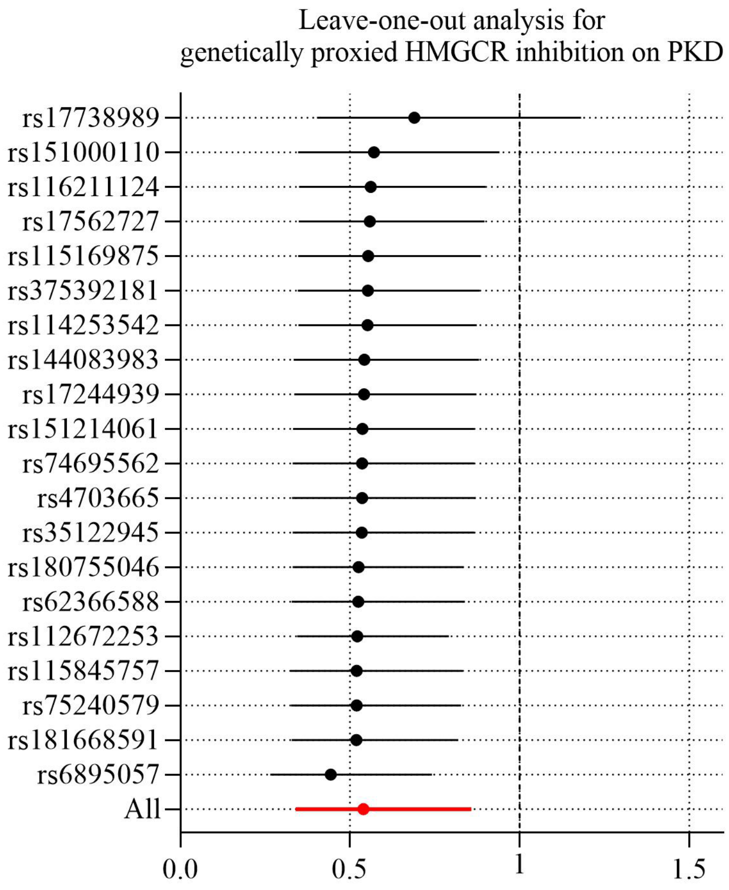

**eFigure 6. Leave-one-out sensitivity analysis for genetically proxied HMGCR inhibition on PKD subtype (TD vs PIGD).**

HMGCR, 3-hydroxy-3-methylglutaryl-CoA reductase; PIGD, postural instability/gait difficulty; PKD, Parkinson's disease; TD, tremor-dominant.

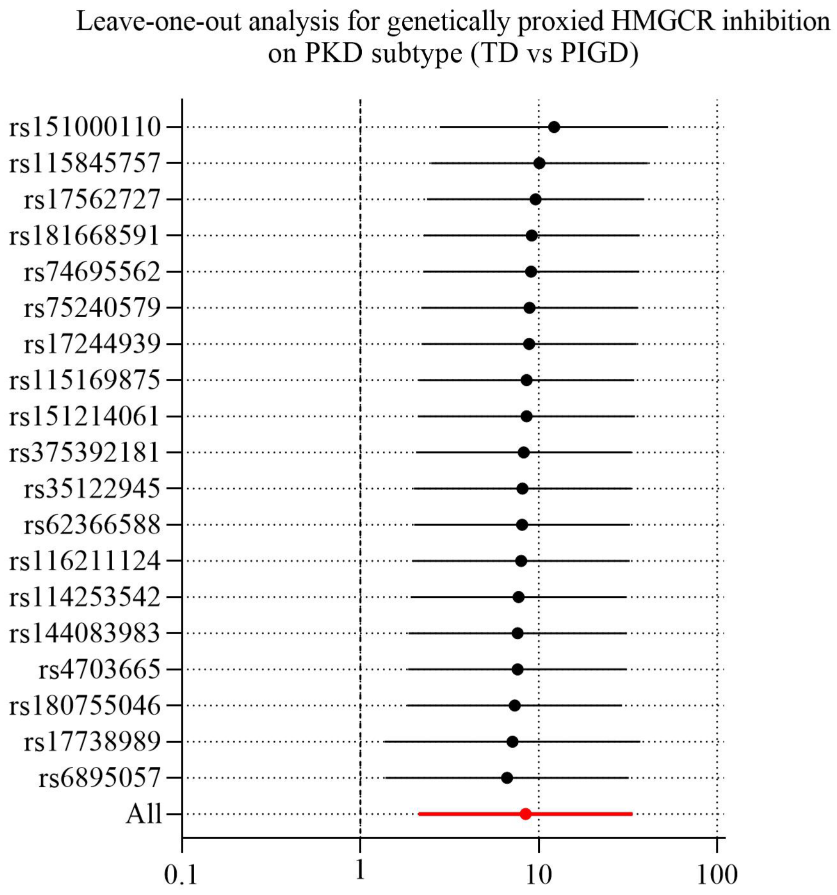

**eFigure 7. Leave-one-out sensitivity analysis for genetically proxied PCSK9 inhibition on PKD subtype (TD vs PIGD).**

PCSK9, proprotein convertase subtilisin/kexin type; PIGD, postural instability/gait difficulty; PKD, Parkinson's disease; TD, tremor-dominant.

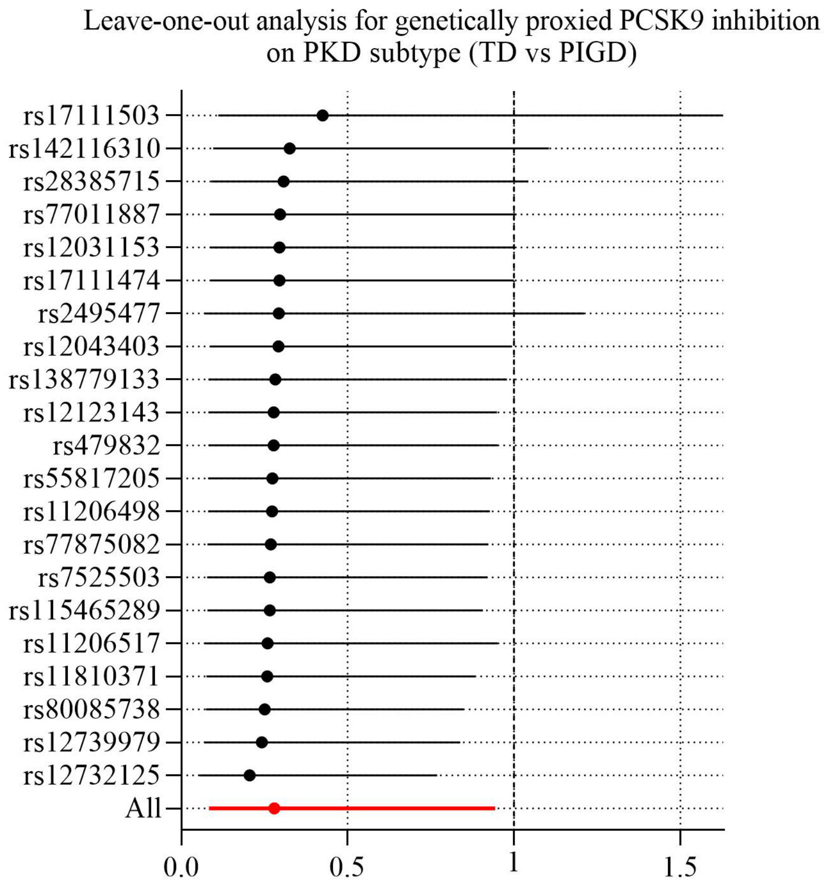

**eTable 1. Genetic variants used to instrument each lipid-lowering drug for the primary analysis.**

EA/OA, effect allele/other allele; MAF, minor allele frequency.

| Target | SNP | EA/OA | MAF | Beta | SE | *P*-value | F |
|---|---|---|---|---|---|---|---|
| HMGCR | rs375392181 | T/G | 0.0101 | 0.0454 | 0.0071 | 1.34E-10 | 41 |
| HMGCR | rs17738989 | T/C | 0.3660 | 0.0475 | 0.0015 | 1.00E-200 | 1044 |
| HMGCR | rs115845757 | A/G | 0.0197 | 0.0580 | 0.0053 | 2.81E-28 | 122 |
| HMGCR | rs4703665 | T/C | 0.1370 | -0.0306 | 0.0023 | 3.27E-41 | 181 |
| HMGCR | rs35122945 | C/A | 0.0674 | -0.0371 | 0.0030 | 1.04E-35 | 156 |
| HMGCR | rs151214061 | C/T | 0.0216 | 0.0429 | 0.0049 | 2.27E-18 | 76 |
| HMGCR | rs6895057 | A/G | 0.2440 | 0.0467 | 0.0016 | 1.72E-185 | 844 |
| HMGCR | rs75240579 | T/C | 0.0401 | -0.0404 | 0.0038 | 5.51E-27 | 116 |
| HMGCR | rs112672253 | T/A | 0.0078 | 0.0576 | 0.0083 | 3.44E-12 | 48 |
| HMGCR | rs17244939 | C/A | 0.0156 | -0.0430 | 0.0062 | 2.84E-12 | 49 |
| HMGCR | rs115169875 | A/G | 0.0257 | -0.0337 | 0.0046 | 2.73E-13 | 53 |
| HMGCR | rs62366588 | A/C | 0.0580 | -0.0346 | 0.0032 | 2.87E-27 | 117 |
| HMGCR | rs74695562 | G/T | 0.0417 | -0.0369 | 0.0037 | 1.80E-23 | 100 |
| HMGCR | rs17562727 | C/T | 0.0274 | 0.0511 | 0.0045 | 2.28E-30 | 131 |
| HMGCR | rs144083983 | T/C | 0.0702 | -0.0418 | 0.0030 | 2.37E-44 | 195 |
| HMGCR | rs181668591 | T/C | 0.0102 | 0.0604 | 0.0072 | 6.53E-17 | 70 |
| HMGCR | rs114253542 | C/T | 0.0074 | 0.0759 | 0.0087 | 1.85E-18 | 77 |
| HMGCR | rs180755046 | A/C | 0.0161 | -0.0421 | 0.0062 | 1.38E-11 | 46 |
| HMGCR | rs182826525 | G/A | 0.0136 | 0.1035 | 0.0086 | 3.73E-33 | 144 |

| | | | | | | | |
|---|---|---|---|---|---|---|---|
| HMGCR | rs116211124 | A/C | 0.0321 | 0.0419 | 0.0040 | 8.80E-26 | 110 |
| HMGCR | rs151000110 | A/G | 0.0566 | 0.0670 | 0.0031 | 1.65E-103 | 467 |
| HMGCR | rs200823803 | C/T | 0.0181 | 0.0551 | 0.0060 | 3.87E-20 | 84 |
| NPC1L1 | rs141530948 | A/G | 0.0062 | -0.0558 | 0.0094 | 2.97E-09 | 35 |
| NPC1L1 | rs77826622 | C/T | 0.0220 | 0.0313 | 0.0049 | 2.26E-10 | 40 |
| NPC1L1 | rs7808295 | T/C | 0.0410 | -0.0231 | 0.0037 | 5.33E-10 | 39 |
| NPC1L1 | rs17725246 | C/T | 0.1950 | 0.0428 | 0.0018 | 2.03E-126 | 572 |
| NPC1L1 | rs77517259 | T/C | 0.0101 | 0.0503 | 0.0073 | 5.10E-12 | 48 |
| NPC1L1 | rs117623941 | T/C | 0.0173 | -0.0315 | 0.0057 | 2.48E-08 | 31 |
| NPC1L1 | rs2289051 | T/C | 0.0259 | 0.0259 | 0.0046 | 1.49E-08 | 32 |
| NPC1L1 | rs217370 | G/A | 0.4670 | -0.0254 | 0.0014 | 4.33E-74 | 332 |
| NPC1L1 | rs79836087 | A/G | 0.0454 | -0.0215 | 0.0034 | 3.74E-10 | 39 |
| NPC1L1 | rs79854399 | T/C | 0.0189 | -0.0390 | 0.0052 | 8.13E-14 | 56 |
| NPC1L1 | rs143116067 | A/G | 0.0675 | -0.0201 | 0.0029 | 8.35E-12 | 47 |
| NPC1L1 | rs2331139 | A/T | 0.0123 | 0.0444 | 0.0079 | 1.91E-08 | 32 |
| PCSK9 | rs12043403 | C/T | 0.0975 | -0.0245 | 0.0025 | 3.01E-23 | 99 |
| PCSK9 | rs12409233 | C/G | 0.0882 | -0.0226 | 0.0024 | 2.50E-20 | 85 |
| PCSK9 | rs11206498 | G/A | 0.3420 | 0.0103 | 0.0015 | 2.21E-12 | 49 |
| PCSK9 | rs890576 | G/C | 0.2020 | -0.0153 | 0.0017 | 2.27E-18 | 76 |
| PCSK9 | rs12123143 | C/T | 0.2290 | 0.0132 | 0.0023 | 5.57E-09 | 34 |
| PCSK9 | rs12732125 | T/C | 0.0188 | -0.1739 | 0.0052 | 1.00E-200 | 1099 |
| PCSK9 | rs145075626 | G/C | 0.0172 | -0.0503 | 0.0055 | 5.98E-20 | 84 |
| PCSK9 | rs17111474 | T/C | 0.2960 | 0.0113 | 0.0016 | 4.50E-12 | 48 |
| PCSK9 | rs77875082 | A/G | 0.0305 | 0.0538 | 0.0042 | 2.75E-37 | 163 |
| PCSK9 | rs12117661 | G/C | 0.2420 | -0.0660 | 0.0016 | 1.00E-200 | 1614 |

| | | | | | | | |
|---|---|---|---|---|---|---|---|
| PCSK9 | rs374459115 | A/G | 0.0172 | 0.0683 | 0.0068 | 5.13E-24 | 102 |
| PCSK9 | rs28775984 | C/T | 0.4320 | -0.0193 | 0.0030 | 2.18E-10 | 40 |
| PCSK9 | rs2479420 | C/T | 0.2610 | 0.0367 | 0.0017 | 6.83E-100 | 450 |
| PCSK9 | rs182491400 | T/C | 0.0209 | 0.0445 | 0.0056 | 1.18E-15 | 64 |
| PCSK9 | rs181331606 | G/C | 0.0901 | 0.0286 | 0.0030 | 6.29E-22 | 93 |
| PCSK9 | rs80085738 | A/G | 0.0109 | -0.0407 | 0.0068 | 2.63E-09 | 35 |
| PCSK9 | rs12739979 | T/C | 0.2380 | -0.0270 | 0.0018 | 2.40E-50 | 223 |
| PCSK9 | rs11810371 | A/G | 0.0428 | -0.0423 | 0.0037 | 9.91E-31 | 133 |
| PCSK9 | rs75050571 | T/A | 0.0236 | 0.0282 | 0.0050 | 1.44E-08 | 32 |
| PCSK9 | rs72660548 | G/C | 0.0202 | 0.0704 | 0.0050 | 6.19E-46 | 202 |
| PCSK9 | rs17111503 | G/A | 0.2560 | 0.0487 | 0.0016 | 1.00E-200 | 920 |
| PCSK9 | rs2479408 | G/C | 0.1990 | -0.0445 | 0.0018 | 2.61E-134 | 608 |
| PCSK9 | rs479832 | C/T | 0.1780 | -0.0144 | 0.0019 | 6.70E-15 | 61 |
| PCSK9 | rs2495477 | G/A | 0.3970 | -0.0524 | 0.0015 | 1.00E-200 | 1234 |
| PCSK9 | rs41294825 | T/A | 0.0430 | -0.0414 | 0.0034 | 7.20E-34 | 147 |
| PCSK9 | rs7525503 | T/G | 0.0229 | 0.0713 | 0.0050 | 2.51E-46 | 204 |
| PCSK9 | rs11206517 | G/T | 0.0371 | 0.0889 | 0.0037 | 1.00E-126 | 573 |
| PCSK9 | rs41297885 | G/C | 0.0358 | -0.0406 | 0.0038 | 7.82E-27 | 115 |
| PCSK9 | rs28385715 | G/T | 0.0229 | 0.0456 | 0.0049 | 1.34E-20 | 87 |
| PCSK9 | rs77011887 | T/C | 0.0170 | 0.0459 | 0.0057 | 8.19E-16 | 65 |
| PCSK9 | rs142116310 | A/G | 0.0107 | 0.0554 | 0.0072 | 1.84E-14 | 59 |
| PCSK9 | rs115465289 | A/G | 0.0312 | -0.0407 | 0.0042 | 6.23E-22 | 93 |
| PCSK9 | rs530804537 | A/G | 0.0121 | -0.2555 | 0.0072 | 1.00E-200 | 1266 |
| PCSK9 | rs12031153 | A/G | 0.0531 | 0.0331 | 0.0035 | 1.07E-21 | 92 |

| | | | | | | | |
|---|---|---|---|---|---|---|---|
| PCSK9 | rs138779133 | G/A | 0.0271 | -0.0599 | 0.0043 | 2.00E-43 | 191 |
| PCSK9 | rs79494709 | C/G | 0.0491 | -0.0258 | 0.0033 | 2.37E-15 | 63 |
| PCSK9 | rs55817205 | A/G | 0.0116 | 0.0676 | 0.0071 | 2.25E-21 | 90 |

**eTable 2. Characteristics of instrumental variables for each drug target in the LDL and hyperlipidemia datasets.**
Chr, chromosome; EA/OA, effect allele/other allele; LDL, low density lipoprotein; Pos, position.

| Drug class | Proxy gene | SNP | Chr | Pos | EA/OA | Exposure (LDL) | | Outcome (hyperlipidemia) | |
|---|---|---|---|---|---|---|---|---|---|
| | | | | | | Beta | SE | Beta | SE |
| HMGCR inhibitors | *HMGCR* | rs112672253 | 5 | 74628706 | T/A | 0.0576 | 0.0083 | -0.0011 | 0.0431 |
| | | rs114253542 | 5 | 74692810 | C/T | 0.0759 | 0.0087 | 0.0499 | 0.0445 |
| | | rs115169875 | 5 | 74633264 | A/G | -0.0337 | 0.0046 | -0.0469 | 0.0232 |
| | | rs115845757 | 5 | 74563700 | A/G | 0.0580 | 0.0053 | 0.0733 | 0.0290 |
| | | rs116211124 | 5 | 74720833 | A/C | 0.0419 | 0.0040 | 0.0403 | 0.0207 |
| | | rs144083983 | 5 | 74683306 | T/C | -0.0418 | 0.0030 | -0.0091 | 0.0145 |
| | | rs151000110 | 5 | 74725216 | A/G | 0.0670 | 0.0031 | 0.0654 | 0.0162 |
| | | rs151214061 | 5 | 74614286 | C/T | 0.0429 | 0.0049 | 0.0493 | 0.0289 |
| | | rs17244939 | 5 | 74631096 | C/A | -0.0430 | 0.0062 | 0.0117 | 0.0266 |
| | | rs17562727 | 5 | 74682474 | C/T | 0.0511 | 0.0045 | 0.0846 | 0.0226 |
| | | rs17738989 | 5 | 74533730 | T/C | 0.0475 | 0.0015 | 0.0451 | 0.0080 |
| | | rs180755046 | 5 | 74703989 | A/C | -0.0421 | 0.0062 | -0.0503 | 0.0271 |
| | | rs181668591 | 5 | 74689045 | T/C | 0.0604 | 0.0072 | 0.0064 | 0.0428 |
| | | rs200823803 | 5 | 74753738 | C/T | 0.0551 | 0.0060 | -0.0268 | 0.0351 |
| | | rs35122945 | 5 | 74610293 | C/A | -0.0371 | 0.0030 | -0.0290 | 0.0156 |
| | | rs375392181 | 5 | 74532691 | T/G | 0.0454 | 0.0071 | 0.0311 | 0.0399 |
| | | rs4703665 | 5 | 74602898 | T/C | -0.0306 | 0.0023 | -0.0355 | 0.0110 |

| | | | | | | | | | |
|---|---|---|---|---|---|---|---|---|---|
| | | rs62366588 | 5 | 74664987 | A/C | -0.0346 | 0.0032 | -0.0194 | 0.0159 |
| | | rs6895057 | 5 | 74624234 | A/G | 0.0467 | 0.0016 | 0.0579 | 0.0089 |
| | | rs74695562 | 5 | 74675951 | G/T | -0.0369 | 0.0037 | -0.0228 | 0.0186 |
| | | rs75240579 | 5 | 74624484 | T/C | -0.0404 | 0.0038 | -0.0179 | 0.0180 |
| NPC1L1 inhibitors | *NPC1L1* | rs117623941 | 7 | 44598725 | T/C | -0.0315 | 0.0057 | -0.0214 | 0.0285 |
| | | rs141530948 | 7 | 44511663 | A/G | -0.0558 | 0.0094 | -0.0494 | 0.0494 |
| | | rs143116067 | 7 | 44653084 | A/G | -0.0201 | 0.0029 | -0.0185 | 0.0155 |
| | | rs17725246 | 7 | 44581986 | C/T | 0.0428 | 0.0018 | 0.0625 | 0.0098 |
| | | rs217370 | 7 | 44615161 | G/A | -0.0254 | 0.0014 | -0.0357 | 0.0077 |
| | | rs2289051 | 7 | 44608718 | T/C | 0.0259 | 0.0046 | 0.0449 | 0.0271 |
| | | rs2331139 | 7 | 44665419 | A/T | 0.0444 | 0.0079 | 0.1317 | 0.0354 |
| | | rs77517259 | 7 | 44586770 | T/C | 0.0503 | 0.0073 | 0.0884 | 0.0471 |
| | | rs77826622 | 7 | 44572346 | C/T | 0.0313 | 0.0049 | 0.0555 | 0.0260 |
| | | rs7808295 | 7 | 44572699 | T/C | -0.0231 | 0.0037 | -0.0048 | 0.0203 |
| | | rs79836087 | 7 | 44635997 | A/G | -0.0215 | 0.0034 | -0.0143 | 0.0185 |
| | | rs79854399 | 7 | 44640315 | T/C | -0.0390 | 0.0052 | -0.0634 | 0.0289 |
| PCSK9 inhibitors | *PCSK9* | rs11206498 | 1 | 55437104 | G/A | 0.0103 | 0.0015 | 0.0080 | 0.0082 |
| | | rs11206517 | 1 | 55526428 | G/T | 0.0889 | 0.0037 | 0.1397 | 0.0205 |
| | | rs115465289 | 1 | 55580914 | A/G | -0.0407 | 0.0042 | -0.0491 | 0.0214 |
| | | rs11810371 | 1 | 55496861 | A/G | -0.0423 | 0.0037 | -0.0744 | 0.0192 |
| | | rs12031153 | 1 | 55601339 | A/G | 0.0331 | 0.0035 | 0.0580 | 0.0170 |
| | | rs12043403 | 1 | 55431933 | C/T | -0.0245 | 0.0025 | -0.0038 | 0.0135 |

| | | | | | | | | |
|---|---|---|---|---|---|---|---|---|
| | rs12117661 | 1 | 55487346 | G/C | -0.0660 | 0.0016 | -0.0858 | 0.0091 |
| | rs12123143 | 1 | 55453856 | C/T | 0.0132 | 0.0023 | -0.0122 | 0.0089 |
| | rs12409233 | 1 | 55436599 | C/G | -0.0226 | 0.0024 | -0.0488 | 0.0135 |
| | rs12732125 | 1 | 55470153 | T/C | -0.1739 | 0.0052 | -0.1517 | 0.0288 |
| | rs12739979 | 1 | 55496648 | T/C | -0.0270 | 0.0018 | -0.0294 | 0.0094 |
| | rs138779133 | 1 | 55604404 | G/A | -0.0599 | 0.0043 | -0.0695 | 0.0248 |
| | rs142116310 | 1 | 55536386 | A/G | 0.0554 | 0.0072 | 0.0718 | 0.0360 |
| | rs145075626 | 1 | 55479043 | G/C | -0.0503 | 0.0055 | -0.0911 | 0.0297 |
| | rs17111474 | 1 | 55479967 | T/C | 0.0113 | 0.0016 | 0.0122 | 0.0087 |
| | rs17111503 | 1 | 55503448 | G/A | 0.0487 | 0.0016 | 0.0784 | 0.0086 |
| | rs181331606 | 1 | 55493672 | G/C | 0.0286 | 0.0030 | 0.0572 | 0.0146 |
| | rs182491400 | 1 | 55493375 | T/C | 0.0445 | 0.0056 | 0.0803 | 0.0266 |
| | rs2479408 | 1 | 55504188 | G/C | -0.0445 | 0.0018 | -0.0646 | 0.0097 |
| | rs2479420 | 1 | 55492190 | C/T | 0.0367 | 0.0017 | 0.0412 | 0.0087 |
| | rs2495477 | 1 | 55518467 | G/A | -0.0524 | 0.0015 | -0.0666 | 0.0081 |
| | rs28385715 | 1 | 55532142 | G/T | 0.0456 | 0.0049 | 0.0627 | 0.0249 |
| | rs28775984 | 1 | 55491161 | C/T | -0.0193 | 0.0030 | -0.0080 | 0.0089 |
| | rs374459115 | 1 | 55489968 | A/G | 0.0683 | 0.0068 | 0.0580 | 0.0343 |
| | rs41294825 | 1 | 55519068 | T/A | -0.0414 | 0.0034 | -0.0477 | 0.0186 |
| | rs41297885 | 1 | 55526840 | G/C | -0.0406 | 0.0038 | -0.0446 | 0.0207 |
| | rs479832 | 1 | 55515537 | C/T | -0.0144 | 0.0019 | -0.0174 | 0.0101 |
| | rs530804537 | 1 | 55583210 | A/G | -0.2555 | 0.0072 | -0.2832 | 0.0413 |

| | | | | | | | | |
|---|---|---|---|---|---|---|---|---|
| | rs55817205 | 1 | 55609056 | A/G | 0.0676 | 0.0071 | 0.1011 | 0.0381 |
| | rs72660548 | 1 | 55500978 | G/C | 0.0704 | 0.0050 | 0.1211 | 0.0273 |
| | rs75050571 | 1 | 55500283 | T/A | 0.0282 | 0.0050 | -0.0403 | 0.0285 |
| | rs7525503 | 1 | 55522558 | T/G | 0.0713 | 0.0050 | 0.1110 | 0.0273 |
| | rs77011887 | 1 | 55534271 | T/C | 0.0459 | 0.0057 | 0.0468 | 0.0274 |
| | rs77875082 | 1 | 55485042 | A/G | 0.0538 | 0.0042 | 0.0582 | 0.0222 |
| | rs79494709 | 1 | 55608984 | C/G | -0.0258 | 0.0033 | -0.0210 | 0.0172 |
| | rs80085738 | 1 | 55495299 | A/G | -0.0407 | 0.0068 | -0.1480 | 0.0374 |
| | rs890576 | 1 | 55452897 | G/C | -0.0153 | 0.0017 | -0.0238 | 0.0096 |

**eTable 3. Characteristics of instrumental variables for each drug target in the LDL and coronary artery disease datasets.** Chr, chromosome; CAD, coronary artery disease; EA/OA, effect allele/other allele; LDL, low density lipoprotein; Pos, position.

| Drug class | Proxy gene | SNP | Chr | Pos | EA/OA | Exposure (LDL) | | Outcome (CAD) | |
|---|---|---|---|---|---|---|---|---|---|
| | | | | | | Beta | SE | Beta | SE |
| HMGCR inhibitors | *HMGCR* | rs112672253 | 5 | 74628706 | T/A | 0.0576 | 0.0083 | 0.0595 | 0.0670 |
| | | rs114253542 | 5 | 74692810 | C/T | 0.0759 | 0.0087 | 0.0829 | 0.0626 |
| | | rs115169875 | 5 | 74633264 | A/G | -0.0337 | 0.0046 | 0.0741 | 0.0459 |
| | | rs115845757 | 5 | 74563700 | A/G | 0.0580 | 0.0053 | 0.0466 | 0.0428 |
| | | rs116211124 | 5 | 74720833 | A/C | 0.0419 | 0.0040 | 0.0428 | 0.0267 |
| | | rs144083983 | 5 | 74683306 | T/C | -0.0418 | 0.0030 | 0.0025 | 0.0281 |
| | | rs151000110 | 5 | 74725216 | A/G | 0.0670 | 0.0031 | 0.0140 | 0.0223 |
| | | rs151214061 | 5 | 74614286 | C/T | 0.0429 | 0.0049 | 0.0107 | 0.0333 |
| | | rs17244939 | 5 | 74631096 | C/A | -0.0430 | 0.0062 | -0.0305 | 0.0593 |
| | | rs17562727 | 5 | 74682474 | C/T | 0.0511 | 0.0045 | 0.0873 | 0.0323 |
| | | rs17738989 | 5 | 74533730 | T/C | 0.0475 | 0.0015 | 0.0071 | 0.0096 |
| | | rs181668591 | 5 | 74689045 | T/C | 0.0604 | 0.0072 | 0.0082 | 0.0654 |
| | | rs35122945 | 5 | 74610293 | C/A | -0.0371 | 0.0030 | -0.0203 | 0.0295 |
| | | rs375392181 | 5 | 74532691 | T/G | 0.0454 | 0.0071 | -0.0478 | 0.0398 |
| | | rs4703665 | 5 | 74602898 | T/C | -0.0306 | 0.0023 | 0.0017 | 0.0168 |
| | | rs62366588 | 5 | 74664987 | A/C | -0.0346 | 0.0032 | -0.0015 | 0.0336 |
| | | rs6895057 | 5 | 74624234 | A/G | 0.0467 | 0.0016 | 0.0198 | 0.0105 |
| | | rs74695562 | 5 | 74675951 | G/T | -0.0369 | 0.0037 | -0.0683 | 0.0289 |

| | | | | | | | | | |
|---|---|---|---|---|---|---|---|---|---|
| | | rs75240579 | 5 | 74624484 | T/C | -0.0404 | 0.0038 | -0.0581 | 0.0426 |
| NPC1L1 inhibitors | *NPC1L1* | rs117623941 | 7 | 44598725 | T/C | -0.0315 | 0.0057 | 0.0051 | 0.0435 |
| | | rs143116067 | 7 | 44653084 | A/G | -0.0201 | 0.0029 | -0.0476 | 0.0236 |
| | | rs17725246 | 7 | 44581986 | C/T | 0.0428 | 0.0018 | 0.0213 | 0.0119 |
| | | rs217370 | 7 | 44615161 | G/A | -0.0254 | 0.0014 | -0.0182 | 0.0093 |
| | | rs2289051 | 7 | 44608718 | T/C | 0.0259 | 0.0046 | 0.0505 | 0.0166 |
| | | rs77517259 | 7 | 44586770 | T/C | 0.0503 | 0.0073 | 0.0222 | 0.0548 |
| | | rs77826622 | 7 | 44572346 | C/T | 0.0313 | 0.0049 | 0.0573 | 0.0458 |
| | | rs7808295 | 7 | 44572699 | T/C | -0.0231 | 0.0037 | -0.0223 | 0.0377 |
| | | rs79836087 | 7 | 44635997 | A/G | -0.0215 | 0.0034 | 0.0053 | 0.0281 |
| | | rs79854399 | 7 | 44640315 | T/C | -0.0390 | 0.0052 | -0.0103 | 0.0392 |
| PCSK9 inhibitors | *PCSK9* | rs11206498 | 1 | 55437104 | G/A | 0.0103 | 0.0015 | -0.0048 | 0.0101 |
| | | rs11206517 | 1 | 55526428 | G/T | 0.0889 | 0.0037 | 0.0500 | 0.0237 |
| | | rs115465289 | 1 | 55580914 | A/G | -0.0407 | 0.0042 | -0.0384 | 0.0354 |
| | | rs11810371 | 1 | 55496861 | A/G | -0.0423 | 0.0037 | 0.0370 | 0.0256 |
| | | rs12031153 | 1 | 55601339 | A/G | 0.0331 | 0.0035 | 0.0409 | 0.0218 |
| | | rs12043403 | 1 | 55431933 | C/T | -0.0245 | 0.0025 | 0.0142 | 0.0193 |
| | | rs12117661 | 1 | 55487346 | G/C | -0.0660 | 0.0016 | -0.0551 | 0.0116 |
| | | rs12123143 | 1 | 55453856 | C/T | 0.0132 | 0.0023 | 0.0122 | 0.0145 |
| | | rs12409233 | 1 | 55436599 | C/G | -0.0226 | 0.0024 | -0.0374 | 0.0187 |
| | | rs12739979 | 1 | 55496648 | T/C | -0.0270 | 0.0018 | -0.0143 | 0.0139 |
| | | rs138779133 | 1 | 55604404 | G/A | -0.0599 | 0.0043 | -0.0554 | 0.0299 |

| | | | | | | | | |
|---|---|---|---|---|---|---|---|---|
| | rs142116310 | 1 | 55536386 | A/G | 0.0554 | 0.0072 | 0.0027 | 0.0699 |
| | rs145075626 | 1 | 55479043 | G/C | -0.0503 | 0.0055 | 0.0089 | 0.0386 |
| | rs17111474 | 1 | 55479967 | T/C | 0.0113 | 0.0016 | 0.0105 | 0.0115 |
| | rs17111503 | 1 | 55503448 | G/A | 0.0487 | 0.0016 | 0.0349 | 0.0113 |
| | rs182491400 | 1 | 55493375 | T/C | 0.0445 | 0.0056 | 0.0304 | 0.0415 |
| | rs2479408 | 1 | 55504188 | G/C | -0.0445 | 0.0018 | -0.0046 | 0.0134 |
| | rs2495477 | 1 | 55518467 | G/A | -0.0524 | 0.0015 | -0.0359 | 0.0105 |
| | rs28385715 | 1 | 55532142 | G/T | 0.0456 | 0.0049 | 0.0554 | 0.0406 |
| | rs41294825 | 1 | 55519068 | T/A | -0.0414 | 0.0034 | -0.0351 | 0.0238 |
| | rs41297885 | 1 | 55526840 | G/C | -0.0406 | 0.0038 | -0.0302 | 0.0259 |
| | rs479832 | 1 | 55515537 | C/T | -0.0144 | 0.0019 | -0.0121 | 0.0131 |
| | rs72660548 | 1 | 55500978 | G/C | 0.0704 | 0.0050 | 0.1096 | 0.0418 |
| | rs75050571 | 1 | 55500283 | T/A | 0.0282 | 0.0050 | -0.0323 | 0.0491 |
| | rs7525503 | 1 | 55522558 | T/G | 0.0713 | 0.0050 | 0.0704 | 0.0293 |
| | rs77875082 | 1 | 55485042 | A/G | 0.0538 | 0.0042 | 0.0122 | 0.0341 |
| | rs79494709 | 1 | 55608984 | C/G | -0.0258 | 0.0033 | -0.0246 | 0.0250 |
| | rs80085738 | 1 | 55495299 | A/G | -0.0407 | 0.0068 | -0.0997 | 0.0622 |
| | rs890576 | 1 | 55452897 | G/C | -0.0153 | 0.0017 | -0.0167 | 0.0113 |

**eTable 4. Characteristics of instrumental variables for each drug target in the LDL and body mass index datasets.**
BMI, body mass index; Chr, chromosome; EA/OA, effect allele/other allele; LDL, low density lipoprotein; Pos, position.

| Drug class | Proxy gene | SNP | Chr | Pos | EA/OA | Exposure (LDL) | | Outcome (BMI) | |
|---|---|---|---|---|---|---|---|---|---|
| | | | | | | Beta | SE | Beta | SE |
| HMGCR inhibitors | *HMGCR* | rs17738989 | 5 | 74533730 | T/C | 0.0475 | 0.0015 | -0.0181 | 0.0018 |
| | | rs4703665 | 5 | 74602898 | T/C | -0.0306 | 0.0023 | 0.0128 | 0.0025 |
| NPC1L1 inhibitors | *NPC1L1* | rs17725246 | 7 | 44581986 | C/T | 0.0428 | 0.0018 | 0.0003 | 0.0021 |
| | | rs217370 | 7 | 44615161 | G/A | -0.0254 | 0.0014 | -0.0002 | 0.0016 |
| | | rs2289051 | 7 | 44608718 | T/C | 0.0259 | 0.0046 | -0.0022 | 0.0051 |
| PCSK9 inhibitors | *PCSK9* | rs11206498 | 1 | 55437104 | G/A | 0.0103 | 0.0015 | 0.0000 | 0.0018 |
| | | rs11206517 | 1 | 55526428 | G/T | 0.0889 | 0.0037 | 0.0011 | 0.0047 |
| | | rs11810371 | 1 | 55496861 | A/G | -0.0423 | 0.0037 | 0.0051 | 0.0045 |
| | | rs12117661 | 1 | 55487346 | G/C | -0.066 | 0.0016 | 0.0014 | 0.0021 |
| | | rs17111503 | 1 | 55503448 | G/A | 0.0487 | 0.0016 | -0.002 | 0.0019 |
| | | rs2479408 | 1 | 55504188 | G/C | -0.0445 | 0.0018 | 0.0011 | 0.0022 |
| | | rs41297885 | 1 | 55526840 | G/C | -0.0406 | 0.0038 | -0.005 | 0.0046 |
| | | rs890576 | 1 | 55452897 | G/C | -0.0153 | 0.0017 | 0.0014 | 0.0021 |

**eTable 5. Characteristics of instrumental variables for each drug target in the LDL and waist circumference datasets.** Chr, chromosome; EA/OA, effect allele/other allele; LDL, low density lipoprotein; Pos, position; WC, waist circumference.

| Drug class | Proxy gene | SNP | Chr | Pos | EA/OA | Exposure (LDL) | | Outcome (WC) | |
|---|---|---|---|---|---|---|---|---|---|
| | | | | | | Beta | SE | Beta | SE |
| HMGCR inhibitors | *HMGCR* | rs112672253 | 5 | 74628706 | T/A | 0.0576 | 0.0083 | -0.0147 | 0.0101 |
| | | rs114253542 | 5 | 74692810 | C/T | 0.0759 | 0.0087 | -0.0064 | 0.0104 |
| | | rs115169875 | 5 | 74633264 | A/G | -0.0337 | 0.0046 | 0.0080 | 0.0055 |
| | | rs115845757 | 5 | 74563700 | A/G | 0.0580 | 0.0053 | -0.0059 | 0.0067 |
| | | rs116211124 | 5 | 74720833 | A/C | 0.0419 | 0.0040 | -0.0223 | 0.0048 |
| | | rs144083983 | 5 | 74683306 | T/C | -0.0418 | 0.0030 | 0.0150 | 0.0034 |
| | | rs151000110 | 5 | 74725216 | A/G | 0.0670 | 0.0031 | -0.0139 | 0.0039 |
| | | rs151214061 | 5 | 74614286 | C/T | 0.0429 | 0.0049 | 0.0157 | 0.0066 |
| | | rs17244939 | 5 | 74631096 | C/A | -0.0430 | 0.0062 | 0.0107 | 0.0062 |
| | | rs17562727 | 5 | 74682474 | C/T | 0.0511 | 0.0045 | 0.0042 | 0.0055 |
| | | rs17738989 | 5 | 74533730 | T/C | 0.0475 | 0.0015 | -0.0170 | 0.0019 |
| | | rs180755046 | 5 | 74703989 | A/C | -0.0421 | 0.0062 | 0.0166 | 0.0064 |
| | | rs181668591 | 5 | 74689045 | T/C | 0.0604 | 0.0072 | -0.0277 | 0.0100 |
| | | rs35122945 | 5 | 74610293 | C/A | -0.0371 | 0.0030 | 0.0041 | 0.0036 |
| | | rs375392181 | 5 | 74532691 | T/G | 0.0454 | 0.0071 | -0.0010 | 0.0090 |
| | | rs4703665 | 5 | 74602898 | T/C | -0.0306 | 0.0023 | 0.0127 | 0.0026 |
| | | rs62366588 | 5 | 74664987 | A/C | -0.0346 | 0.0032 | 0.0163 | 0.0037 |
| | | rs6895057 | 5 | 74624234 | A/G | 0.0467 | 0.0016 | -0.0111 | 0.0021 |

| | | | | | | | | | |
|---|---|---|---|---|---|---|---|---|---|
| | | rs74695562 | 5 | 74675951 | G/T | -0.0369 | 0.0037 | -0.0141 | 0.0043 |
| | | rs75240579 | 5 | 74624484 | T/C | -0.0404 | 0.0038 | 0.0024 | 0.0042 |
| NPC1L1 inhibitors | *NPC1L1* | rs117623941 | 7 | 44598725 | T/C | -0.0315 | 0.0057 | -0.0019 | 0.0067 |
| | | rs141530948 | 7 | 44511663 | A/G | -0.0558 | 0.0094 | 0.0061 | 0.0116 |
| | | rs143116067 | 7 | 44653084 | A/G | -0.0201 | 0.0029 | 0.0018 | 0.0036 |
| | | rs17725246 | 7 | 44581986 | C/T | 0.0428 | 0.0018 | 0.0011 | 0.0023 |
| | | rs217370 | 7 | 44615161 | G/A | -0.0254 | 0.0014 | -0.0061 | 0.0018 |
| | | rs2289051 | 7 | 44608718 | T/C | 0.0259 | 0.0046 | -0.0034 | 0.0062 |
| | | rs77517259 | 7 | 44586770 | T/C | 0.0503 | 0.0073 | 0.0124 | 0.011 |
| | | rs77826622 | 7 | 44572346 | C/T | 0.0313 | 0.0049 | -0.0125 | 0.0061 |
| | | rs7808295 | 7 | 44572699 | T/C | -0.0231 | 0.0037 | -0.0032 | 0.0048 |
| | | rs79836087 | 7 | 44635997 | A/G | -0.0215 | 0.0034 | -0.0111 | 0.0042 |
| | | rs79854399 | 7 | 44640315 | T/C | -0.0390 | 0.0052 | 0.0002 | 0.0067 |
| PCSK9 inhibitors | *PCSK9* | rs11206498 | 1 | 55437104 | G/A | 0.0103 | 0.0015 | -0.0012 | 0.0019 |
| | | rs11206517 | 1 | 55526428 | G/T | 0.0889 | 0.0037 | 0.0002 | 0.0050 |
| | | rs115465289 | 1 | 55580914 | A/G | -0.0407 | 0.0042 | 0.0072 | 0.0050 |
| | | rs11810371 | 1 | 55496861 | A/G | -0.0423 | 0.0037 | 0.0016 | 0.0044 |
| | | rs12031153 | 1 | 55601339 | A/G | 0.0331 | 0.0035 | -0.0054 | 0.0040 |
| | | rs12043403 | 1 | 55431933 | C/T | -0.0245 | 0.0025 | 0.0037 | 0.0031 |
| | | rs12117661 | 1 | 55487346 | G/C | -0.066 | 0.0016 | 0.0017 | 0.0021 |
| | | rs12123143 | 1 | 55453856 | C/T | 0.0132 | 0.0023 | -0.0009 | 0.0021 |
| | | rs12409233 | 1 | 55436599 | C/G | -0.0226 | 0.0024 | 0.0009 | 0.0031 |

| | | | | | | | | |
|---|---|---|---|---|---|---|---|---|
| | rs12732125 | 1 | 55470153 | T/C | -0.1739 | 0.0052 | 0.0020 | 0.0063 |
| | rs12739979 | 1 | 55496648 | T/C | -0.027 | 0.0018 | 0.0016 | 0.0022 |
| | rs138779133 | 1 | 55604404 | G/A | -0.0599 | 0.0043 | 0.0041 | 0.0058 |
| | rs142116310 | 1 | 55536386 | A/G | 0.0554 | 0.0072 | -0.0051 | 0.0084 |
| | rs145075626 | 1 | 55479043 | G/C | -0.0503 | 0.0055 | 0.0042 | 0.0069 |
| | rs17111474 | 1 | 55479967 | T/C | 0.0113 | 0.0016 | -0.0029 | 0.0020 |
| | rs17111503 | 1 | 55503448 | G/A | 0.0487 | 0.0016 | -0.0021 | 0.0020 |
| | rs2479408 | 1 | 55504188 | G/C | -0.0445 | 0.0018 | 0.0015 | 0.0022 |
| | rs2495477 | 1 | 55518467 | G/A | -0.0524 | 0.0015 | 0.0022 | 0.0019 |
| | rs28385715 | 1 | 55532142 | G/T | 0.0456 | 0.0049 | 0.0143 | 0.0058 |
| | rs41294825 | 1 | 55519068 | T/A | -0.0414 | 0.0034 | 0.0008 | 0.0043 |
| | rs41297885 | 1 | 55526840 | G/C | -0.0406 | 0.0038 | 0.0070 | 0.0049 |
| | rs479832 | 1 | 55515537 | C/T | -0.0144 | 0.0019 | 0.0039 | 0.0023 |
| | rs55817205 | 1 | 55609056 | A/G | 0.0676 | 0.0071 | 0.0029 | 0.0089 |
| | rs72660548 | 1 | 55500978 | G/C | 0.0704 | 0.0050 | 0.0069 | 0.0067 |
| | rs75050571 | 1 | 55500283 | T/A | 0.0282 | 0.0050 | -0.0031 | 0.0065 |
| | rs7525503 | 1 | 55522558 | T/G | 0.0713 | 0.0050 | 0.0060 | 0.0065 |
| | rs77011887 | 1 | 55534271 | T/C | 0.0459 | 0.0057 | 0.0020 | 0.0064 |
| | rs77875082 | 1 | 55485042 | A/G | 0.0538 | 0.0042 | 0.0020 | 0.0052 |
| | rs79494709 | 1 | 55608984 | C/G | -0.0258 | 0.0033 | 0.0032 | 0.004 |
| | rs80085738 | 1 | 55495299 | A/G | -0.0407 | 0.0068 | -0.0041 | 0.0083 |
| | rs890576 | 1 | 55452897 | G/C | -0.0153 | 0.0017 | 0.0030 | 0.0022 |

**eTable 6. Characteristics of instrumental variables for each drug target in the LDL and Hip circumference datasets.** Chr, chromosome; EA/OA, effect allele/other allele; HC, hip circumference; LDL, low density lipoprotein; Pos, position.

| Drug class | Proxy gene | SNP | Chr | Pos | EA/OA | Exposure (LDL) | | Outcome (HC) | |
|---|---|---|---|---|---|---|---|---|---|
| | | | | | | Beta | SE | Beta | SE |
| HMGCR inhibitors | *HMGCR* | rs112672253 | 5 | 74628706 | T/A | 0.0576 | 0.0083 | -0.0227 | 0.0112 |
| | | rs114253542 | 5 | 74692810 | C/T | 0.0759 | 0.0087 | -0.0102 | 0.0116 |
| | | rs115169875 | 5 | 74633264 | A/G | -0.0337 | 0.0046 | 0.0101 | 0.0061 |
| | | rs115845757 | 5 | 74563700 | A/G | 0.0580 | 0.0053 | -0.0011 | 0.0075 |
| | | rs116211124 | 5 | 74720833 | A/C | 0.0419 | 0.0040 | -0.0256 | 0.0054 |
| | | rs144083983 | 5 | 74683306 | T/C | -0.0418 | 0.0030 | 0.0138 | 0.0038 |
| | | rs151000110 | 5 | 74725216 | A/G | 0.0670 | 0.0031 | -0.0208 | 0.0043 |
| | | rs151214061 | 5 | 74614286 | C/T | 0.0429 | 0.0049 | 0.0199 | 0.0073 |
| | | rs17244939 | 5 | 74631096 | C/A | -0.0430 | 0.0062 | 0.0077 | 0.0069 |
| | | rs17562727 | 5 | 74682474 | C/T | 0.0511 | 0.0045 | -0.0035 | 0.0061 |
| | | rs17738989 | 5 | 74533730 | T/C | 0.0475 | 0.0015 | -0.0188 | 0.0021 |
| | | rs180755046 | 5 | 74703989 | A/C | -0.0421 | 0.0062 | 0.0169 | 0.0071 |
| | | rs181668591 | 5 | 74689045 | T/C | 0.0604 | 0.0072 | -0.0156 | 0.0112 |
| | | rs35122945 | 5 | 74610293 | C/A | -0.0371 | 0.0030 | 0.0042 | 0.0040 |
| | | rs375392181 | 5 | 74532691 | T/G | 0.0454 | 0.0071 | -0.0072 | 0.0101 |
| | | rs4703665 | 5 | 74602898 | T/C | -0.0306 | 0.0023 | 0.0144 | 0.0028 |
| | | rs62366588 | 5 | 74664987 | A/C | -0.0346 | 0.0032 | 0.0185 | 0.0041 |
| | | rs6895057 | 5 | 74624234 | A/G | 0.0467 | 0.0016 | -0.0146 | 0.0023 |

| | | | | | | | | | |
|---|---|---|---|---|---|---|---|---|---|
| | | rs74695562 | 5 | 74675951 | G/T | -0.0369 | 0.0037 | -0.0089 | 0.0048 |
| | | rs75240579 | 5 | 74624484 | T/C | -0.0404 | 0.0038 | 0.0055 | 0.0047 |
| NPC1L1 inhibitors | *NPC1L1* | rs117623941 | 7 | 44598725 | T/C | -0.0315 | 0.0057 | -0.0069 | 0.0074 |
| | | rs141530948 | 7 | 44511663 | A/G | -0.0558 | 0.0094 | 0.0164 | 0.0130 |
| | | rs143116067 | 7 | 44653084 | A/G | -0.0201 | 0.0029 | 0.0037 | 0.004 |
| | | rs17725246 | 7 | 44581986 | C/T | 0.0428 | 0.0018 | 0.0011 | 0.0026 |
| | | rs217370 | 7 | 44615161 | G/A | -0.0254 | 0.0014 | -0.0065 | 0.0020 |
| | | rs2289051 | 7 | 44608718 | T/C | 0.0259 | 0.0046 | -0.0113 | 0.0069 |
| | | rs77517259 | 7 | 44586770 | T/C | 0.0503 | 0.0073 | 0.0090 | 0.0123 |
| | | rs77826622 | 7 | 44572346 | C/T | 0.0313 | 0.0049 | -0.0097 | 0.0068 |
| | | rs7808295 | 7 | 44572699 | T/C | -0.0231 | 0.0037 | -0.0030 | 0.0053 |
| | | rs79836087 | 7 | 44635997 | A/G | -0.0215 | 0.0034 | -0.0121 | 0.0047 |
| | | rs79854399 | 7 | 44640315 | T/C | -0.0390 | 0.0052 | 0.0033 | 0.0075 |
| PCSK9 inhibitors | *PCSK9* | rs11206498 | 1 | 55437104 | G/A | 0.0103 | 0.0015 | -0.0014 | 0.0021 |
| | | rs11206517 | 1 | 55526428 | G/T | 0.0889 | 0.0037 | 0.0065 | 0.0055 |
| | | rs115465289 | 1 | 55580914 | A/G | -0.0407 | 0.0042 | 0.0129 | 0.0055 |
| | | rs11810371 | 1 | 55496861 | A/G | -0.0423 | 0.0037 | 0.0037 | 0.0049 |
| | | rs12031153 | 1 | 55601339 | A/G | 0.0331 | 0.0035 | -0.0061 | 0.0045 |
| | | rs12043403 | 1 | 55431933 | C/T | -0.0245 | 0.0025 | 0.0080 | 0.0035 |
| | | rs12117661 | 1 | 55487346 | G/C | -0.0660 | 0.0016 | -0.0003 | 0.0023 |
| | | rs12123143 | 1 | 55453856 | C/T | 0.0132 | 0.0023 | -0.0004 | 0.0023 |
| | | rs12409233 | 1 | 55436599 | C/G | -0.0226 | 0.0024 | -0.0006 | 0.0034 |

| | | | | | | | | |
|---|---|---|---|---|---|---|---|---|
| | rs12732125 | 1 | 55470153 | T/C | -0.1739 | 0.0052 | -0.0018 | 0.0070 |
| | rs12739979 | 1 | 55496648 | T/C | -0.0270 | 0.0018 | -0.0017 | 0.0024 |
| | rs138779133 | 1 | 55604404 | G/A | -0.0599 | 0.0043 | 0.0041 | 0.0064 |
| | rs142116310 | 1 | 55536386 | A/G | 0.0554 | 0.0072 | -0.0009 | 0.0094 |
| | rs145075626 | 1 | 55479043 | G/C | -0.0503 | 0.0055 | -0.0051 | 0.0077 |
| | rs17111474 | 1 | 55479967 | T/C | 0.0113 | 0.0016 | -0.0018 | 0.0022 |
| | rs17111503 | 1 | 55503448 | G/A | 0.0487 | 0.0016 | -0.0011 | 0.0023 |
| | rs2479408 | 1 | 55504188 | G/C | -0.0445 | 0.0018 | -0.0013 | 0.0025 |
| | rs2495477 | 1 | 55518467 | G/A | -0.0524 | 0.0015 | -0.0005 | 0.0021 |
| | rs28385715 | 1 | 55532142 | G/T | 0.0456 | 0.0049 | 0.0066 | 0.0065 |
| | rs41294825 | 1 | 55519068 | T/A | -0.0414 | 0.0034 | 0.0029 | 0.0048 |
| | rs41297885 | 1 | 55526840 | G/C | -0.0406 | 0.0038 | 0.0048 | 0.0054 |
| | rs479832 | 1 | 55515537 | C/T | -0.0144 | 0.0019 | 0.0034 | 0.0026 |
| | rs55817205 | 1 | 55609056 | A/G | 0.0676 | 0.0071 | 0.0071 | 0.0099 |
| | rs72660548 | 1 | 55500978 | G/C | 0.0704 | 0.0050 | 0.0071 | 0.0074 |
| | rs75050571 | 1 | 55500283 | T/A | 0.0282 | 0.0050 | 0.0017 | 0.0073 |
| | rs7525503 | 1 | 55522558 | T/G | 0.0713 | 0.0050 | 0.0075 | 0.0073 |
| | rs77011887 | 1 | 55534271 | T/C | 0.0459 | 0.0057 | 0.0026 | 0.0072 |
| | rs77875082 | 1 | 55485042 | A/G | 0.0538 | 0.0042 | 0.0021 | 0.0058 |
| | rs79494709 | 1 | 55608984 | C/G | -0.0258 | 0.0033 | 0.0005 | 0.0045 |
| | rs80085738 | 1 | 55495299 | A/G | -0.0407 | 0.0068 | -0.0021 | 0.0092 |
| | rs890576 | 1 | 55452897 | G/C | -0.0153 | 0.0017 | 0.0030 | 0.0025 |

**eTable 7. Characteristics of instrumental variables for HMGCR protein target in the LDL and Parkinson's disease datasets.**
Chr, chromosome; EA/OA, effect allele/other allele; LDL, low density lipoprotein; PIGD, postural instability/gait difficulty; PKD, Parkinson's disease; Pos, position; TD, tremor-dominant.

| SNP | Chr | Pos | EA/OA | Exposure (LDL) | | Outcome (PKD) | | Outcome (TD vs PIGD) | |
|---|---|---|---|---|---|---|---|---|---|
| | | | | Beta | SE | Beta | SE | Beta | SE |
| rs112672253 | 5 | 74628706 | T/A | 0.0576 | 0.0083 | -0.4005 | 0.1830 | — | — |
| rs114253542 | 5 | 74692810 | C/T | 0.0759 | 0.0087 | 0.3014 | 0.1961 | -0.5122 | 0.3852 |
| rs115169875 | 5 | 74633264 | A/G | -0.0337 | 0.0046 | -0.0875 | 0.0616 | 0.0315 | 0.2181 |
| rs115845757 | 5 | 74563700 | A/G | 0.0580 | 0.0053 | -0.0414 | 0.0741 | 0.2716 | 0.2552 |
| rs116211124 | 5 | 74720833 | A/C | 0.0419 | 0.0040 | 0.0799 | 0.0513 | -0.1656 | 0.1693 |
| rs144083983 | 5 | 74683306 | T/C | -0.0418 | 0.0030 | -0.0316 | 0.0470 | 0.1958 | 0.1499 |
| rs151000110 | 5 | 74725216 | A/G | 0.0670 | 0.0031 | 0.0719 | 0.0422 | 0.0319 | 0.1343 |
| rs151214061 | 5 | 74614286 | C/T | 0.0429 | 0.0049 | 0.0127 | 0.0709 | -0.0645 | 0.2100 |

| | | | | | | | | | |
|---|---|---|---|---|---|---|---|---|---|
| rs17244939 | 5 | 74631096 | C/A | -0.0430 | 0.0062 | -0.0440 | 0.1001 | -0.1076 | 0.3109 |
| rs17562727 | 5 | 74682474 | C/T | 0.0511 | 0.0045 | 0.0961 | 0.0650 | 0.1034 | 0.2082 |
| rs17738989 | 5 | 74533730 | T/C | 0.0475 | 0.0015 | 0.0548 | 0.0178 | -0.1200 | 0.0612 |
| rs180755046 | 5 | 74703989 | A/C | -0.0421 | 0.0062 | 0.0863 | 0.0903 | 0.9569 | 0.3606 |
| rs181668591 | 5 | 74689045 | T/C | 0.0604 | 0.0072 | -0.1157 | 0.1032 | 0.1349 | 0.3182 |
| rs35122945 | 5 | 74610293 | C/A | -0.0371 | 0.0030 | -0.0113 | 0.0501 | 0.1291 | 0.1541 |
| rs375392181 | 5 | 74532691 | T/G | 0.0454 | 0.0071 | 0.1232 | 0.0887 | -0.1832 | 0.2986 |
| rs4703665 | 5 | 74602898 | T/C | -0.0306 | 0.0023 | -0.0141 | 0.0303 | 0.1356 | 0.1036 |
| rs62366588 | 5 | 74664987 | A/C | -0.0346 | 0.0032 | 0.0370 | 0.0572 | 0.1693 | 0.1914 |
| rs6895057 | 5 | 74624234 | A/G | 0.0467 | 0.0016 | 0.0000 | 0.0200 | -0.1378 | 0.0692 |
| rs74695562 | 5 | 74675951 | G/T | -0.0369 | 0.0037 | -0.0117 | 0.0511 | -0.0428 | 0.1765 |

| | | | | | | | | | |
|---|---|---|---|---|---|---|---|---|---|
| rs75240579 | 5 | 74624484 | T/C | -0.0404 | 0.0038 | 0.0479 | 0.0595 | -0.0074 | 0.1961 |

**eTable 8. Characteristics of instrumental variables for NPC1L1 protein target in the LDL and Parkinson's disease datasets.**
Chr, chromosome; EA/OA, effect allele/other allele; LDL, low density lipoprotein; PIGD, postural instability/gait difficulty; PKD, Parkinson's disease; Pos, position; TD, tremor-dominant.

| SNP | Chr | Pos | EA/OA | Exposure (LDL) | | Outcome (PKD) | | Outcome (TD vs PIGD) | |
|---|---|---|---|---|---|---|---|---|---|
| | | | | Beta | SE | Beta | SE | Beta | SE |
| rs117623941 | 7 | 44598725 | T/C | -0.0315 | 0.0057 | 0.0591 | 0.0966 | 0.3590 | 0.2408 |
| rs141530948 | 7 | 44511663 | A/G | -0.0558 | 0.0094 | 0.0527 | 0.1744 | 0.6263 | 0.5320 |
| rs143116067 | 7 | 44653084 | A/G | -0.0201 | 0.0029 | 0.0211 | 0.0483 | 0.2568 | 0.1318 |
| rs17725246 | 7 | 44581986 | C/T | 0.0428 | 0.0018 | 0.0392 | 0.0232 | -0.0794 | 0.0808 |
| rs217370 | 7 | 44615161 | G/A | -0.0254 | 0.0014 | 0.0067 | 0.0203 | 0.0697 | 0.0626 |
| rs2289051 | 7 | 44608718 | T/C | 0.0259 | 0.0046 | 0.0192 | 0.0588 | -0.2920 | 0.2187 |
| rs77517259 | 7 | 44586770 | T/C | 0.0503 | 0.0073 | 0.1299 | 0.1767 | 0.2051 | 0.4055 |
| rs77826622 | 7 | 44572346 | C/T | 0.0313 | 0.0049 | 0.1354 | 0.0800 | 0.0789 | 0.2581 |

| | | | | | | | | | |
|---|---|---|---|---|---|---|---|---|---|
| rs7808295 | 7 | 44572699 | T/C | -0.0231 | 0.0037 | -0.0163 | 0.0752 | -0.2341 | 0.1838 |
| rs79836087 | 7 | 44635997 | A/G | -0.0215 | 0.0034 | -0.0418 | 0.0539 | 0.0998 | 0.1639 |
| rs79854399 | 7 | 44640315 | T/C | -0.0390 | 0.0052 | -0.0206 | 0.0816 | -0.0743 | 0.2729 |

**eTable 9. Characteristics of instrumental variables for PCSK9 protein target in the LDL and Parkinson’s disease datasets.**
Chr, chromosome; EA/OA, effect allele/other allele; LDL, low density lipoprotein; PIGD, postural instability/gait difficulty; PKD, Parkinson’s disease; Pos, position; TD, tremor-dominant.

| SNP | Chr | Pos | EA/OA | Exposure (LDL) | | Outcome (PKD) | | Outcome (TD vs PIGD) | |
|---|---|---|---|---|---|---|---|---|---|
| | | | | Beta | SE | Beta | SE | Beta | SE |
| rs11206498 | 1 | 55437104 | G/A | 0.0103 | 0.0015 | -0.0062 | 0.0236 | -0.0146 | 0.0698 |
| rs11206517 | 1 | 55526428 | G/T | 0.0889 | 0.0037 | -0.0340 | 0.0464 | 0.0642 | 0.1587 |
| rs115465289 | 1 | 55580914 | A/G | -0.0407 | 0.0042 | 0.0064 | 0.0690 | 0.0685 | 0.1963 |
| rs11810371 | 1 | 55496861 | A/G | -0.0423 | 0.0037 | -0.0656 | 0.0616 | 0.0990 | 0.1813 |
| rs12031153 | 1 | 55601339 | A/G | 0.0331 | 0.0035 | 0.0051 | 0.0558 | 0.1961 | 0.1904 |
| rs12043403 | 1 | 55431933 | C/T | -0.0245 | 0.0025 | — | — | -0.0988 | 0.1221 |
| rs12117661 | 1 | 55487346 | G/C | -0.0660 | 0.0016 | 0.0275 | 0.0216 | — | — |
| rs12123143 | 1 | 55453856 | C/T | 0.0132 | 0.0023 | -0.0469 | 0.0345 | 0.0087 | 0.0871 |

| | | | | | | | | | |
|---|---|---|---|---|---|---|---|---|---|
| rs12409233 | 1 | 55436599 | C/G | -0.0226 | 0.0024 | -0.0023 | 0.0385 | 0.0442 | 0.1111 |
| rs12732125 | 1 | 55470153 | T/C | -0.1739 | 0.0052 | 0.0796 | 0.0849 | 0.0767 | 0.2761 |
| rs12739979 | 1 | 55496648 | T/C | -0.0270 | 0.0018 | 0.0203 | 0.0311 | 0.0530 | 0.0813 |
| rs138779133 | 1 | 55604404 | G/A | -0.0599 | 0.0043 | -0.0053 | 0.0711 | -0.0962 | 0.2077 |
| rs142116310 | 1 | 55536386 | A/G | 0.0554 | 0.0072 | 0.3727 | 0.1734 | 0.7585 | 0.3130 |
| rs145075626 | 1 | 55479043 | G/C | -0.0503 | 0.0055 | -0.1014 | 0.0944 | — | — |
| rs17111474 | 1 | 55479967 | T/C | 0.0113 | 0.0016 | 0.0316 | 0.0311 | 0.0912 | 0.0793 |
| rs17111503 | 1 | 55503448 | G/A | 0.0487 | 0.0016 | 0.0249 | 0.0206 | 0.1551 | 0.0712 |
| rs2479408 | 1 | 55504188 | G/C | -0.0445 | 0.0018 | -0.0033 | 0.0252 | — | — |
| rs2495477 | 1 | 55518467 | G/A | -0.0524 | 0.0015 | -0.0254 | 0.0233 | -0.0742 | 0.0642 |
| rs28385715 | 1 | 55532142 | G/T | 0.0456 | 0.0049 | -0.0427 | 0.0759 | 0.3927 | 0.2515 |

| | | | | | | | | | |
|---|---|---|---|---|---|---|---|---|---|
| rs41294825 | 1 | 55519068 | T/A | -0.0414 | 0.0034 | 0.0001 | 0.0554 | — | — |
| rs41297885 | 1 | 55526840 | G/C | -0.0406 | 0.0038 | 0.0055 | 0.0496 | — | — |
| rs479832 | 1 | 55515537 | C/T | -0.0144 | 0.0019 | -0.0346 | 0.0257 | -0.0116 | 0.0783 |
| rs55817205 | 1 | 55609056 | A/G | 0.0676 | 0.0071 | -0.0899 | 0.1791 | -0.0749 | 0.4444 |
| rs72660548 | 1 | 55500978 | G/C | 0.0704 | 0.0050 | -0.0084 | 0.0832 | — | — |
| rs7525503 | 1 | 55522558 | T/G | 0.0713 | 0.0050 | 0.0169 | 0.0675 | -0.0024 | 0.2322 |
| rs77011887 | 1 | 55534271 | T/C | 0.0459 | 0.0057 | — | — | 0.3547 | 0.2974 |
| rs77875082 | 1 | 55485042 | A/G | 0.0538 | 0.0042 | -0.0565 | 0.0685 | -0.0115 | 0.2087 |
| rs79494709 | 1 | 55608984 | C/G | -0.0258 | 0.0033 | -0.0498 | 0.0444 | — | — |
| rs80085738 | 1 | 55495299 | A/G | -0.0407 | 0.0068 | — | — | 0.5008 | 0.2805 |
| rs890576 | 1 | 55452897 | G/C | -0.0153 | 0.0017 | 0.0166 | 0.0276 | — | — |

**eTable 10. Colocalization results in the target gene region for LDL and PKD.**
The region was defined as ±100kb base pairs of gene region.

Colocalization analysis essentially examines the posterior probability of hypothesis H4 (both traits are associated and share a single causal variant). In statistics, a higher posterior probability for hypothesis H4 indicates a better explanation of how significant signal loci impact the phenotype. The range of H4 value is between 0 and 1, with a higher posterior probability considered more favorable. In general literature, loci with posterior probabilities between 0.75 and 0.95 are considered colocalized loci. However, there is an inherent distinction between colocalization analysis and Mendelian Randomization (MR).The purpose of colocalization analysis is to investigate whether two traits have a common causal variant, whereas MR analysis aims to explore the association between two traits using genetic variation as instrumental variables. Therefore, colocalization may impose relatively stringent requirements on the correlation between variant trait 1 and variant trait 2, while MR analysis only requires a robust association between the instrumental variable and the exposure.

| Drug class | Drug target encoding gene | H0 | H1 | H2 | H3 | H4 |
|---|---|---|---|---|---|---|
| HMGCR inhibitors | *HMGCR* | 0.00% | 93.80% | 0.00% | 5.34% | 0.87% |

Posterior probability for
H0: neither trait has a genetic association in the region;
H1: only LDL has a genetic association in the region;
H2: only PKD has a genetic association in the region;
H3: both traits are associated, but with different causal variants;
H4: both traits are associated and share a single causal variant.

**eTable 11. Colocalization results in the target gene region for LDL and PKD subtype (TD vs PIGD).**

The region was defined as ±100kb base pairs of gene region.

| Drug class | Drug target encoding gene | H0 | H1 | H2 | H3 | H4 |
|---|---|---|---|---|---|---|
| HMGCR inhibitors | *HMGCR* | 0.00% | 83.60% | 0.00% | 11.00% | 5.31% |
| PCSK9 inhibitors | *PCSK9* | 0.00% | 88.20% | 0.00% | 3.57% | 8.22% |

Posterior probability for

H0: neither trait has a genetic association in the region;

H1: only LDL has a genetic association in the region;

H2: only PKD subtype (TD vs PIGD) has a genetic association in the region;

H3: both traits are associated, but with different causal variants;

H4: both traits are associated and share a single causal variant.
